# Methamphetamine use reshapes cellular and plasma immune signatures in a lineage-specific manner in people with HIV

**DOI:** 10.64898/2026.11.19.26368492

**Authors:** Jonathon P. Sens, Ashish Kumar, Sheri L. Towe, Sun H. Park, Fang-Chi Hsu, Kip D. Zimmerman, Ashish K. Khanna, Kiran K. Solingapuram Sai, E. Alfonso Romero-Sandoval, Gagan Deep, Christina S. Meade, Drew D. Kiraly

**Author notes:** Co-Corresponding authors: To Whom Correspondence Should be Addressed: Christina S. Meade, PhD Professor, Translational Neuroscience, Wake Forest University School of Medicine, 575 N. Patterson Ave., Winston-Salem, NC 27101, Drew D. Kiraly, MD, PhD Associate Professor, Translational Neuroscience and Psychiatry Wake Forest University School of Medicine, 115 South Chestnut St., Winston-Salem, NC 27103.

## Abstract

Methamphetamine use is common among people with HIV (PWH) and is associated with worse neurocognitive and clinical outcomes, but how it reshapes HIV-associated immune dysregulation across cell types and compartments is unknown. In this cross-sectional and observational study, we performed single-cell RNA sequencing of peripheral blood mononuclear cells (PBMCs) from controls without HIV, PWH without methamphetamine use and PWH who use methamphetamine, then scored pathway enrichment, cytokine response and transcription factor activity in each cell population. We then profiled plasma from the same three groups in a larger set of participants by targeted proteomics, with PWH who use cocaine added as a stimulant comparison. In the single-cell data, methamphetamine use acted in opposite directions across lineages. HIV-associated changes were reversed most often in monocytes and CD8^+^ T cells, with interferon, antigen presentation and JAK-STAT programs shifting in monocytes and IFN-γ, IL-2 and MYC programs shifting in CD8^+^ T cells, whereas HIV-associated proliferative and TNF signaling programs persisted in CD4^+^ and γδ T cells of Meth+HIV participants. Central memory CD8^+^ T cells of Meth+HIV participants showed reduced type 1 effector programs. In plasma, HIV was associated with a cytotoxic lymphocyte activation signature (KLRK1, IFN-γ, TNF, granzyme B and CXCL9) that was further elevated with methamphetamine use, together with higher IL-6, CXCL13 and neurofilament heavy chain. IFN-γ was the most connected protein in the plasma interaction network, linking a cytotoxic module to a lymphoid chemokine module. These data define lineage- and compartment-specific immune consequences of methamphetamine use in PWH and nominate IFN-γ signaling, acting through monocyte antigen presentation and CD8^+^ T cell effector programs, together with TNF, IL-6 and CXCL13, as candidate mediators of neuroimmune dysfunction in PWH who use methamphetamine.

**Highlights:**

- Paired single-cell PBMC and NULISA proteomic profiling in PWH who use meth
- HIV-linked T cell activation persists with meth, while monocyte IFN programs flip
- Meth reshapes CD8^+^ T cell interferon and effector programs in people with HIV
- Plasma cytotoxic and neuronal injury markers rise further with meth use
- Monocyte antigen presentation programs suppressed in HIV rebound with meth use

## Introduction

Methamphetamine use has resurged sharply in the United States. Approximately 2.4 million Americans reported past-year methamphetamine use in 2024, and stimulant-involved overdose deaths have tripled over the past decade^1–3^. Methamphetamine use is especially common among people with HIV (PWH), with estimated rates roughly seven times those of the general population^1,4^, and among sexual and gender minorities, persistent methamphetamine use accounts for a large share of new HIV infections^5^. For PWH, the consequences extend well beyond transmission risk, as methamphetamine use is associated with accelerated disease progression, poorer adherence to antiretroviral therapy (ART) and worse neurocognitive outcomes^6–8^. Between 53% and 58% of PWH who use methamphetamine show neurocognitive impairment, compared with approximately 40% of those with either condition alone, with additive deficits in attention, executive function and decision-making^7,9–11^. The biological mechanisms underlying these compounding effects remain poorly understood, and no targeted interventions are approved.

Beyond its effects on plasma viral load, HIV infection produces broad and persistent immunological consequences, even under sustained viral suppression. ART controls viremia, but chronic immune activation persists, characterized by dysregulated inflammatory signaling and altered myeloid and lymphoid cell function^12,13^. Among the most durable features of this state is the expansion of CD8^+^ T lymphocytes, which does not normalize over time, even with viral suppression^14–16^. Activated CD8^+^ T cells accumulate in the cerebrospinal fluid (CSF) and brain during acute and chronic infection^17–19^, and ongoing T cell activation in CSF is associated with neurocognitive decline^20^. CSF CD8^+^ T cells that express interferon-γ (IFN-γ) correlate with the severity of HIV-associated neurocognitive disorders, and IFN-γ-producing, non-cytolytic CD8^+^ T cells appear to drive pathology through chronic inflammatory signaling, whereas cytolytic CD8^+^ T cells help control viral replication^21^. How the balance between these CD8^+^ T cell states is determined in circulating cells, and how substance use alters this balance, has not been defined in PWH.

Methamphetamine alters this immune landscape through several mechanisms. In mice with chronic viral infection, methamphetamine increases PD-1 expression and chemokine receptors associated with brain trafficking on CD8^+^ T cells and impairs T cell subset progression and polyfunctionality^22,23^. In PWH on suppressive ART, methamphetamine use alters T cell function and promotes CD8^+^ T cell proliferation across memory subsets^24^, and cumulative stimulant exposure is associated with adverse immune function outcomes^25^. In SIV-infected macaques, methamphetamine increases inflammatory and Toll-like receptor signaling programs and reprograms gene expression in brain microglia toward cellular profiles linked to inflammation, susceptibility to viral infection, altered metabolism, and cell death^26–28^. Methamphetamine also disrupts the blood-brain barrier through oxidative stress and tight junction degradation, increasing peripheral leukocyte entry into the central nervous system (CNS)^29,30^. Mechanistic attention in neuroHIV has largely focused on myeloid cells, particularly microglia and monocyte-derived macrophages^31–37^, whereas substance use neuroscience has rarely considered adaptive immunity. We recently showed that CD8^+^ T cells recruited to mesolimbic structures by the chemokine CXCL13 are required for opioid self-administration^38^, adding to evidence that CXCL13 and CD8^+^ T cells can act directly on neural function^39–43^.

Despite these advances, how methamphetamine use reshapes the immune landscape of PWH across cell types and compartments has not been systematically defined. Most human studies have only measured a limited set of circulating markers or a single immune population. This leaves many questions unanswered such as which effects of methamphetamine compound those of HIV, which oppose them, and whether peripheral changes are accompanied by circulating markers of CNS injury. Here, we combined single-cell RNA sequencing of peripheral blood mononuclear cells (PBMCs) from controls without HIV, PWH without methamphetamine use, and PWH who use methamphetamine with pathway, cytokine response, transcription factor and protein interaction analyses. We paired these data with targeted plasma proteomics, including a panel of neurological and CNS-related proteins, in a cohort that also included PWH who use cocaine. We find that methamphetamine use leads to lineage-specific alterations. HIV-associated activation programs persist in CD4^+^ and γδ T cells, whereas methamphetamine use shifts interferon and antigen presentation programs in monocytes, and IFN-γ, IL-2 and MYC programs in CD8^+^ T cells, in the opposite direction of HIV infection. The monocyte changes we identify span the antigen presentation machinery and parallel interferon signatures in CD8^+^ T cells. Within the CD8^+^ T cell compartment, central memory cells of Meth+HIV participants show reduced type 1 effector programs. In plasma, HIV is associated with a rise in neurofilament heavy chain and a cytotoxic lymphocyte activation signature that is further elevated with methamphetamine use.

## Materials & Methods

### Study participants and design

Peripheral blood samples were obtained from adults participating in research examining the effects of HIV and stimulant use on neuroimmune and neurobehavioral functioning. The study sample consisted of PWH who used methamphetamine in the past month (Meth+HIV), along with comparison groups of PWH without methamphetamine use (HIV) and people without HIV (Control). Participant characteristics are summarized in Table 1. HIV diagnosis was confirmed through electronic health records. In addition, participants with HIV had to be on stable ART and have plasma HIV RNA levels <200 copies/mL quantified via PCR testing on the day of the blood draw. For participants without HIV, a rapid HIV-1/2 antibody test (OraQuick ADVANCE®) was conducted using a droplet of whole blood collected by pinprick. Participants in the Meth+HIV group reported at least weekly use of methamphetamine in the past month and had a positive urine drug screen for methamphetamine. Participants in the HIV and Control groups reported 0 days of methamphetamine use in the past 12 months, no history of regular methamphetamine use, and no lifetime stimulant use disorder. For plasma proteomics, PWH who used cocaine (Coc+HIV) were also included, with eligibility criteria that mirrored those for the Meth+HIV group, with cocaine substituted as the primary drug of use. Nicotine, alcohol, and marijuana use was permissible in all groups. Exclusion criteria included limited English proficiency or inability to read, severe neurological disease or injury (other than HIV-related infections), acute psychiatric symptoms, autoimmune or hematological disorders affecting immune function, organ or bone marrow transplantation, dialysis, and receipt of immunomodulatory therapies (e.g., chemotherapy, radiation, blood transfusions). Study procedures were approved by the Wake Forest Health Sciences IRB, and all participants provided written informed consent prior to completing any study activities.

**Table 1.** Demographic and clinical characteristics of participants profiled by single-cell RNA sequencing.

|  | **Control**<br>**N = 4** | **HIV**<br>**N = 4** | **Meth+HIV**<br>**N = 4** |
|---|---|---|---|
| Age in years, M ± SD (range) | 45.00 ± 10.23 (35-58) | 43.25 ± 9.22 (36-56) | 44.00 ± 11.28 (33-58) |
| Male sex at birth, n (%) | 4 (100%) | 4 (100%) | 4 (100%) |
| Race, White : Black | 2:2 | 2:2 | 2:2 |
| Education in years, M ± SD | 15.75 ± 2.06 | 18.25 ± 1.71 | 14.50 ± 3.42 |
| Years since HIV diagnosis, M ± SD |  | 13.34 ± 3.52 | 12.36 ± 8.08 |
| Nadir CD4 count, M ± SD |  | 230.50 ± 126.86 |  |
| Current CD4 T-cell count, M ± SD |  | 706.00 ± 200.76 | 507.00 ± 278.35 |
| Current CD8 T-cell count, M ± SD |  | 1099.25 ± 429.50 | 555.75 ± 266.84 |
| Years of regular meth use, M ± SD |  |  | 6.5 ± 1.00 |
| Days of meth use in past 30, M ± SD |  |  | 22.25 ± 8.06 |

Values are mean ± SD (range) for age and mean ± SD for other continuous variables; blank cells, not applicable or not collected.

### Blood collection and PBMC isolation

Non-fasting blood samples were collected by peripheral venipuncture. Plasma for proteomic analyses was obtained from EDTA-treated blood samples using standard processing procedures, aliquoted, and stored at −80°C until assay. For transcriptomic analyses, peripheral blood mononuclear cells (PBMCs) were isolated from Cell Preparation Tube (CPT) vacutainers processed within 2 hours of collection. CPT tubes were centrifuged at 1,800 × g for 30 minutes at 20°C, and the PBMC fraction was harvested, washed twice in phosphate-buffered saline (PBS) by centrifugation at 300 × g for 15 minutes, and assessed for cell concentration and viability using trypan blue exclusion and automated cell counting. PBMCs were cryopreserved in fetal bovine serum containing 10% dimethyl sulfoxide (DMSO), aliquoted at approximately 5 × 10⁵ cells per vial, frozen at −80°C using a controlled-rate freezing method, and transferred to liquid nitrogen storage within 3 days of processing. Cryopreserved PBMCs remained in liquid nitrogen until RNA extraction and sequencing.

### Single-cell RNA sequencing

PBMCs (n = 4 per group) were processed on the Parse Biosciences Evercode WT platform. PBMCs were fixed and permeabilized with the Evercode Cell Fixation kit (ECFC3300) following the protocol of the manufacturer, then underwent split-pool combinatorial barcoding with the Evercode Whole Transcriptome kit v3 (ECWT3300) to multiplex samples^44^. Libraries were prepared as specified by the manufacturer, checked on a BioAnalyzer 2100 (Agilent) and a Qubit fluorometer (Thermo Fisher), and sequenced on an Illumina NovaSeq X Plus (2 × 100 bp). Reads were aligned to hg38 and demultiplexed with split-pipe v1.5.1 (Parse Biosciences).

### Single-cell data processing, quality control and cell type annotation

Gene expression matrices were processed and analyzed in R with Seurat v5.4.0^45^. To exclude low-quality cells and potential multiplets, we removed cells with fewer than 300 or more than 7,000 unique genes, fewer than 1,000 or more than 50,000 unique RNA transcripts, or more than 10% mitochondrial gene expression. Expression values of the remaining cells were normalized with SCTransform^46^ and clustered on a shared nearest neighbor graph, with UMAP for visualization. After quality control, 42,140 cells were carried into secondary analysis, with a mean of 94,000 reads per cell. Cell identities were assigned from the expression of canonical lineage markers, with *MS4A1*, *CD79A* and *CD74* for B cells, *LYZ*, *LST1* and *FCN1* for monocytes, *FCRL6*, *PDGFD* and *KLRD1* for NK cells, *TRDC* for γδ T cells, and *CD3E*, *CD4*, *IL7R*, *CCR7*, *CD8A*, *CD8B* and *NKG7* for CD4^+^ and CD8^+^ T cells. CD8^+^ T cell subcluster identities were assigned from marker gene expression, with *LTB*, *IL7R*, *SELL*, *CCR7*, *TCF7*, *MAL* and *NRCAM* for naive and central memory CD8^+^ T cells, *NKG7*, *PRF1* and *GNLY* for cytotoxic effector CD8^+^ T cells, *GZMK* and *IFNG-AS1* for GZMK⁺ memory CD8^+^ T cells, and *CX3CR1*, *FCRL6* and *TBX21* for CX3CR1⁺ effector memory CD8^+^ T cells.

### Pseudobulk differential expression

For each cell type, we summed counts across all cells from each participant (AggregateExpression) and tested differential expression with DESeq2 through the Seurat FindMarkers function^47^. This method statistically controls for differences in cell counts across samples. A Pseudobulk approach was selected because they demonstrate reasonable control of the type 1 error rates, without the computational burden and failure to converge problems associated with linear mixed models and smaller sample sizes. We computed differential expression in six PBMC populations (CD4^+^ T, CD8^+^ T, γδ T, B and NK cells and monocytes) and five CD8^+^ T cell subtypes (naive, central memory, GZMK⁺ memory, CX3CR1⁺ effector memory and cytotoxic effector), with four participants per group. Groups were Control (people without HIV), HIV (PWH without methamphetamine use) and Meth+HIV (PWH who use methamphetamine). We tested three contrasts, HIV vs Control (the HIV effect), Meth+HIV vs HIV (the effect of methamphetamine use within HIV) and Meth+HIV vs Control (the net effect).

### Gene set enrichment analysis

Genes were ranked by sign(log2 fold change) × −log10(*p*), and preranked GSEA^48^ was run in gseapy^49^ (1,000 permutations; gene sets of 15 to 500 genes) against the MSigDB^50^ Hallmark, KEGG, Reactome, PID, BioCarta, GO and ImmuneSigDB collections. For cross-contrast summaries, we used 44 Hallmark and selected KEGG pathways per cell type, after excluding Hallmark sets specific to non-immune tissues. Each pathway was classified as reversed (FDR < 0.25 in both the HIV and methamphetamine contrasts, in opposite directions), amplified (FDR < 0.25 in both, same direction), changed with HIV only, changed with methamphetamine only, or unchanged. Antigen presentation gene sets in the GO, KEGG, Reactome, BioCarta and PID collections were identified by name (antigen processing, antigen presentation, MHC, peptide antigen or cross-presentation, excluding antigen receptor sets). For candidate monocyte to CD8^+^ T cell contacts, canonical ligand-receptor pairs for antigen presentation, adhesion, co-stimulation, SLAM family, cytokine, inhibitory, and chemokine signaling were retained when the ligand was detected in monocytes and the receptor in CD8^+^ T cells (at least 75% of pseudobulk samples).

### Cytokine response, pathway activity and transcription factor activity

We estimated cytokine response (CytoSig^51^), signaling pathway (PROGENy^52^) and transcription factor activity (DoRothEA regulons, confidence levels A to C^53^) from the differential expression results for each cell type and contrast. Each resource provides, for every cytokine, pathway or transcription factor, a set of target genes with weights that give the direction and strength of each gene response when that signal is active. For each signature, we fit a univariate linear model^54^ in which the gene-level statistic used for GSEA (sign of the log2 fold change multiplied by −log10 *p*) was regressed on the signature weights across all measured target genes. The activity score is the t statistic of the slope, the slope divided by its standard error, so it reflects both how strongly and how consistently the expression changes match the signature. A positive score indicates that genes induced by the signal were higher, and genes repressed by it lower, in the first group of the contrast, and a negative score indicates the reverse. Signatures with fewer than 10 measured target genes were not scored. *p* values from the t distribution were adjusted by the Benjamini-Hochberg method^55^ within each resource, cell type and contrast, and FDR < 0.05 was considered significant. These scores infer signaling activity from transcriptional responses and do not measure cytokine or transcription factor levels directly. Cytokine responses are shown only for cell types in which the receptor genes were detected (each receptor chain in at least 75% of pseudobulk samples).

### Sensitivity analyses

To test whether results were driven by ribosomal, mitochondrial or proliferation genes, we repeated GSEA and activity scoring after removing ribosomal protein and mitochondrially encoded genes, and repeated activity scoring after adjusting for membership in the Hallmark G2M checkpoint and E2F target gene sets. We also repeated pathway classification at FDR < 0.05.

### Protein interaction networks

Interaction networks were built from STRING v12.5^56^. For PBMC populations, we used leading-edge genes from the top Hallmark pathways in each cell type with differential expression *p* < 0.1. For proteomic data, we used proteins with an HIV- or methamphetamine-associated change. The STRING combined score threshold was set for each network (0.40 to 0.85) and is shown in each panel.

### Plasma proteomics using NULISAseq

Plasma proteins were measured with the NULISAseq Inflammation Panel 250 and CNS Disease Panel 120 (Alamar Biosciences, Fremont, CA, USA) on the Alamar ARGO HT platform, following the protocol of the manufacturer and as described previously^57,58^. Plasma samples were thawed overnight at 4 °C and centrifuged at 2,000 × g for 5 minutes, and 25 µL of plasma per sample was used for each panel. Each plate included three replicates of a sample control to assess the intra-plate coefficient of variation, three replicates of an inter-plate control (pooled plasma) for normalization, and four negative controls to estimate the limit of detection (LOD). Target proteins in each sample were bound by a cocktail of capture and detection antibodies linked to DNA barcodes. The immunocomplexes were captured and washed on magnetic oligo-dT beads, released into a low-salt buffer and captured again on streptavidin beads, and the proximal ends of the DNA strands on each immunocomplex were ligated to generate a reporter molecule containing both target-and sample-specific barcodes. Reporter molecules were pooled, amplified by PCR and purified. The pooled library was sequenced on an Illumina NextSeq 2000 using a P2 flow cell. FASTQ files were uploaded to Argo Command Center for normalization. Target counts in each well were divided by the internal control counts of that well and then by the target-specific median of the inter-plate control replicates on the same plate, and the normalized counts were rescaled by adding 1 and log2-transformed to give NULISA Protein Quantification (NPQ) units. Differences in NPQ can therefore be interpreted as log2 fold changes. The LOD for each target was calculated as the mean plus three standard deviations of the normalized counts in the negative control wells, and detectability was defined as the percentage of samples above the LOD.

The Inflammation and CNS panels quantified 249 and 131 proteins, respectively^59^. Samples came from controls without HIV (n = 12), PWH without methamphetamine use (n = 12) and PWH who use methamphetamine (n = 5). PWH who use cocaine (n = 6) were assayed alongside these groups for comparison. Values of zero were treated as missing, and principal component analysis identified no outlier samples.

Groups were compared with Welch t tests^60^, and *p* values were adjusted by the Benjamini-Hochberg method within each panel and comparison. The primary comparisons were HIV vs Control, Meth+HIV vs HIV and Meth+HIV vs Control, and the same comparisons were made for the cocaine group. On the basis of nominal *p* < 0.05, proteins were grouped by whether an HIV-associated change was amplified, reversed, persisted or was not seen with methamphetamine use, or whether they changed with methamphetamine use only; full definitions are given in the Figure 7 legend. A methamphetamine-associated change was considered shared with cocaine when the matching cocaine comparison changed in the same direction at *p* < 0.05.

Selected proteins were compared across Control, HIV and Meth+HIV with pairwise Welch t tests, with *p* values adjusted for the three comparisons by the Holm-Šídák method^61,62^.

### Statistical analysis and software

Single-cell data were processed in R with Seurat 5.4.0, and downstream analyses were run in Python 3.13 with gseapy, SciPy and networkx. All tests were two-sided.

### Data and code availability

Raw sequencing data have been deposited in the Gene Expression Omnibus and will be publicly available upon publication. All other data supporting the findings of this study are available from the corresponding author upon reasonable request, subject to applicable institutional material transfer agreements.

## Results

### Study design and analytic approach

We studied three groups, people without HIV (Control), PWH without methamphetamine use (HIV) and PWH who use methamphetamine (Meth+HIV) (**Fig. 1**; **Table 1**). PBMCs from four participants per group were profiled by single-cell RNA sequencing. The three groups were similar in age (mean 43 to 45 years) and racial composition (two White and two Black participants per group), and all participants were assigned male sex at birth (**Table 1**). Participants in the HIV group had been diagnosed a mean of 13.3 years earlier and had a mean current CD4^+^ T cell count of 706 cells/µL, Meth+HIV participants had been diagnosed a mean of 12.4 years earlier and had a mean current CD4^+^ T cell count of 507 cells/µL, and Meth+HIV participants reported a mean of 6.5 years of regular methamphetamine use and use on 22 of the past 30 days. Plasma from a larger set of participants (Control, n = 12; HIV, n = 12; Meth+HIV, n = 5) was profiled by NULISAseq, together with PWH who use cocaine (Coc+HIV, n = 6) as a stimulant comparison group. The cohort did not include people without HIV who use methamphetamine, so throughout we treat HIV vs Control as the HIV effect and Meth+HIV vs HIV as the effect of methamphetamine use in the context of HIV. Comparing Meth+HIV participants with controls (the Net contrast) shows the combined state of PWH who use methamphetamine.

**Figure 1.**
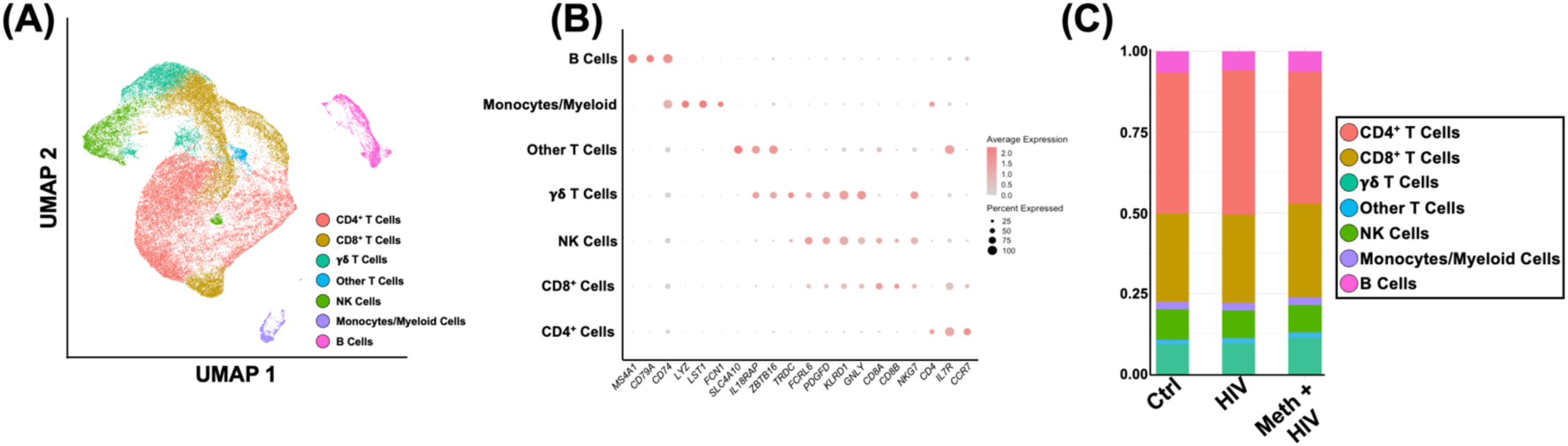
Single-cell RNA sequencing identifies the major PBMC populations in Control, HIV and Meth+HIV participants. Single-cell RNA sequencing of PBMCs from Control, HIV and Meth+HIV participants (n = 4 per group), with 42,140 cells retained after quality control. **(A)** UMAP of all PBMCs, colored by annotated cell type (CD4^+^ T cells, CD8^+^ T cells, γδ T cells, other T cells, NK cells, monocytes and other myeloid cells, and B cells). **(B)** Expression of canonical lineage marker genes across the seven populations. Dot color, average expression; dot size, percentage of cells in the population expressing the gene. **(C)** Proportion of each population in Control, HIV and Meth+HIV participants. Each proportion was compared across groups by one-way ANOVA with no significant differences.

### Methamphetamine use opposes HIV-associated changes in monocytes and CD8^+^ T cells but not in CD4^+^ and γδ T cells

After quality control filtering, 42,140 cells from 12 PBMC samples (n = 4 per group) were retained for analysis. Unsupervised clustering, UMAP visualization and canonical marker gene expression identified the major immune populations expected in human PBMCs, including CD4^+^ T cells, CD8^+^ T cells, γδ T cells, other T cell populations, natural killer (NK) cells, monocytes and other myeloid cells, and B cells (**Fig. 1A, B**). Cell type proportions differed modestly across groups, but none of these differences were significant, indicating that neither HIV nor methamphetamine use substantially altered the overall composition of circulating PBMC populations in this cohort (**Fig. 1C**).

Across the six PBMC populations, we classified 44 Hallmark and selected KEGG pathways per cell type by whether methamphetamine use reversed, had no additional effect on, or added to HIV-associated changes (FDR < 0.25; Supplementary Fig. S1). Methamphetamine use opposed HIV-associated changes most often in monocytes (12 of 19 altered pathways reversed) and CD8^+^ T cells (5 of 12), whereas HIV-only changes predominated in CD4^+^ and γδ T cells, and methamphetamine use did not amplify an HIV-associated pathway change in any cell type. These patterns held at FDR < 0.05 and after removal of ribosomal and mitochondrial genes from the ranked gene lists (Supplementary Figs. S2, S3).

### HIV-associated proliferative and TNF programs persist in CD4^+^ and γδ T cells but are reversed in CD8^+^ T cells

Across T cell populations, HIV was associated with enrichment of growth and translation programs (**Fig. 2A**). In CD4^+^ T cells, HIV enriched cell cycle (E2F targets and G2M checkpoint), spliceosome, MYC target, ribosome and TNFα signaling via NF-κB programs and depleted TCR signaling through PLCγ and IP3R (**2A – Left**; all FDR < 0.01). γδ T cells showed HIV-associated enrichment of MYC targets, ribosome and spliceosome (**2A – Middle**). CD8^+^ T cells shared the MYC target, ribosome and spliceosome enrichment but also showed HIV-associated depletion of IFN-γ response, IFN-α response and allograft rejection programs (**2A – Right**).

**Figure 2.**
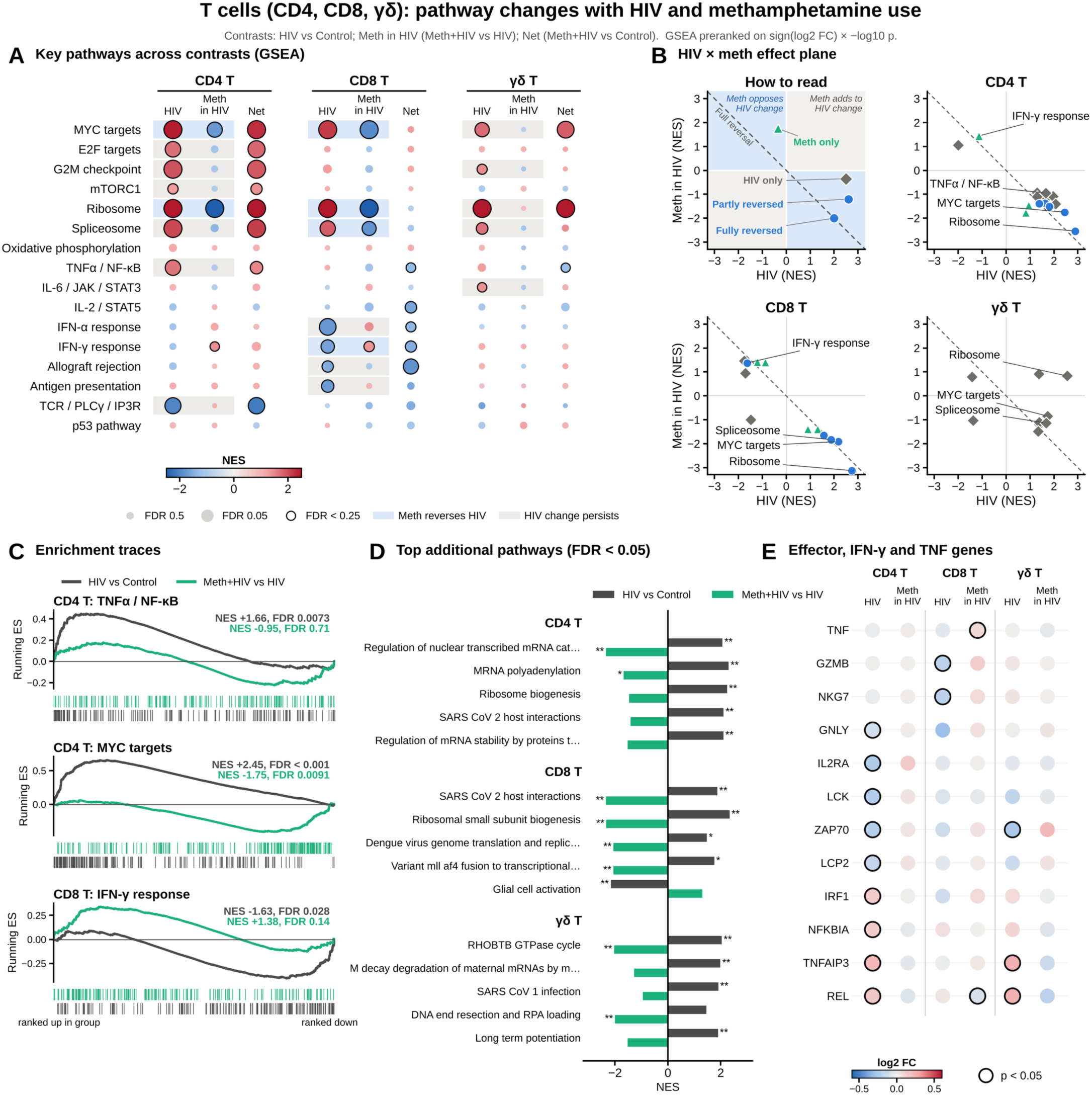
HIV-associated proliferative and TNF programs persist in CD4^+^ and γδ T cells but are reversed in Meth+HIV CD8^+^ T cells. GSEA of gene expression changes in CD4, CD8^+^ and γδ T cells (n = 4 per group). Contrasts: HIV, HIV vs Control; Meth in HIV, Meth+HIV vs HIV; Net, Meth+HIV vs Control. **(A)** NES for 16 key Hallmark and KEGG pathways in each contrast. **(B)** Effect plane relating the effects of HIV and of methamphetamine use within HIV on each pathway. Top left, key: x, NES in the HIV contrast; y, NES in the Meth in HIV contrast; blue quadrants, methamphetamine use moves a pathway in the opposite direction to HIV; gray quadrants, same direction; dashed line, full reversal (y = −x); example points show a fully reversed pathway (on the dashed line), a partly reversed pathway (between the dashed line and the x axis), a pathway changed with HIV only and a pathway changed with methamphetamine use only. Other panels, pathways changed at FDR < 0.25 in either contrast in CD4, CD8^+^ and γδ T cells, plotted by NES. Blue circles, reversed; green triangles, Meth in HIV only; gray diamonds, HIV only. Dashed line, full reversal (y = −x). **(C)** Running enrichment scores for CD4^+^ T cell TNFα signaling via NF-κB, CD4^+^ T cell MYC targets and CD8^+^ T cell IFN-γ response, with genes ranked from most increased (left) to most decreased (right), for the HIV (dark gray) and Meth in HIV (green) contrasts. **(D)** Additional pathways beyond the 16 shown in panel A. For each cell type, gene sets from Reactome, KEGG, GO Biological Process and PID with FDR < 0.05 in either contrast were ranked by FDR. * FDR < 0.25, ** FDR < 0.05. **(E)** Pseudobulk expression changes in the HIV and Meth in HIV contrasts for the 12 genes, from a predefined list of effector, IFN-γ, TNF and TCR signaling genes, with the smallest *p* value in either contrast across the three cell types.

To compare the two effects directly, we plotted each significantly changed pathway by its NES in the HIV contrast (x axis) and the methamphetamine contrast (y axis), with the dashed diagonal marking reversal (**Fig. 2B**). The first panel of **Fig. 2B** shows interpretation of this graph. Pathways on the diagonal are fully reversed, pathways between the diagonal and the x axis are partly reversed, and pathways in the gray quadrants change in the same direction with both exposures. We used this to demonstrate changes in the three T cell populations. In CD4^+^ T cells, the cell cycle, spliceosome, TNFα/NF-κB and TCR signaling pathways were HIV-only changes and remained altered in Meth+HIV participants relative to controls. MYC targets and ribosome were reversed but lay between the diagonal and the x axis, indicating an effect of methamphetamine use within HIV that was smaller than the HIV effect, and both remained enriched relative to controls. In CD8^+^ T cells, MYC targets, ribosome and spliceosome lay close to the diagonal, reflecting reversal of similar magnitude to the HIV effect (for example, MYC targets, HIV NES = 2.17; Meth in HIV NES = −1.92; both FDR < 0.001), so that these programs no longer differed from controls in Meth+HIV participants. IFN-γ response was also reversed in CD8^+^ T cells, whereas IFN-α response and allograft rejection changed with HIV only. In γδ T cells, all nine altered pathways were HIV-only changes.

Running enrichment scores show these patterns for individual pathways (**Fig. 2C**). For TNFα/NF-κB in CD4^+^ T cells, the HIV trace peaked among the most up-regulated genes (NES = 1.66, FDR = 0.007), whereas the methamphetamine trace remained near zero (NES = −0.95, FDR = 0.71). For MYC targets in CD4^+^ T cells, the methamphetamine trace was inverted relative to the HIV trace but smaller in magnitude (NES = −1.75 and 2.45). For IFN-γ response in CD8^+^ T cells, the two traces ran in opposite directions (HIV NES = −1.63, FDR = 0.028; Meth in HIV NES = 1.38, FDR = 0.14).

Beyond the 16 pathways in panel A, we searched the larger Reactome, KEGG, GO Biological Process and PID collections and show, for each cell type, the five most significant gene sets that did not duplicate a panel A pathway (**Fig. 2D**). These were dominated by translation machinery. CD4^+^ T cells showed HIV-associated enrichment of mRNA processing and ribosome biogenesis gene sets, and in CD8^+^ T cells ribosomal small subunit biogenesis was enriched with HIV and depleted with methamphetamine use. To link these pathway changes to individual genes, we examined a predefined set of effector, IFN-γ, TNF and TCR signaling genes and display the 12 most changed across the three T cell populations (**Fig. 2E**). CD4^+^ T cells from PWH expressed less of the proximal TCR signaling genes *LCK*, *ZAP70* and *LCP2* and more of the NF-κB genes *NFKBIA*, *REL* and *TNFAIP3*, and CD8^+^ T cells expressed less of the cytotoxic genes *GZMB* and *NKG7*.

### Methamphetamine use reverses HIV-associated suppression of interferon and antigen presentation programs in monocytes

In B cells, NK cells and monocytes, HIV was associated with opposite patterns of pathway enrichment between lymphocytes and monocytes (**Fig. 3A**). In B cells, HIV was associated with strong enrichment of ribosome (NES = 2.43, FDR < 0.001) and, more modestly, MYC target, spliceosome and TNFα signaling via NF-κB programs (**Fig. 3A, left**). NK cells showed a similar but broader growth signature, with HIV-associated enrichment of ribosome, MYC targets and spliceosome together with the cell cycle programs E2F targets and G2M checkpoint (**Fig. 3A, middle**). Monocytes showed the opposite direction of change, with HIV-associated depletion of IFN-γ response, IFN-α response, allograft rejection, antigen processing and presentation, IL-2/STAT5 signaling and ribosome programs (**Fig. 3A, right;** all FDR < 0.05).

**Figure 3.**
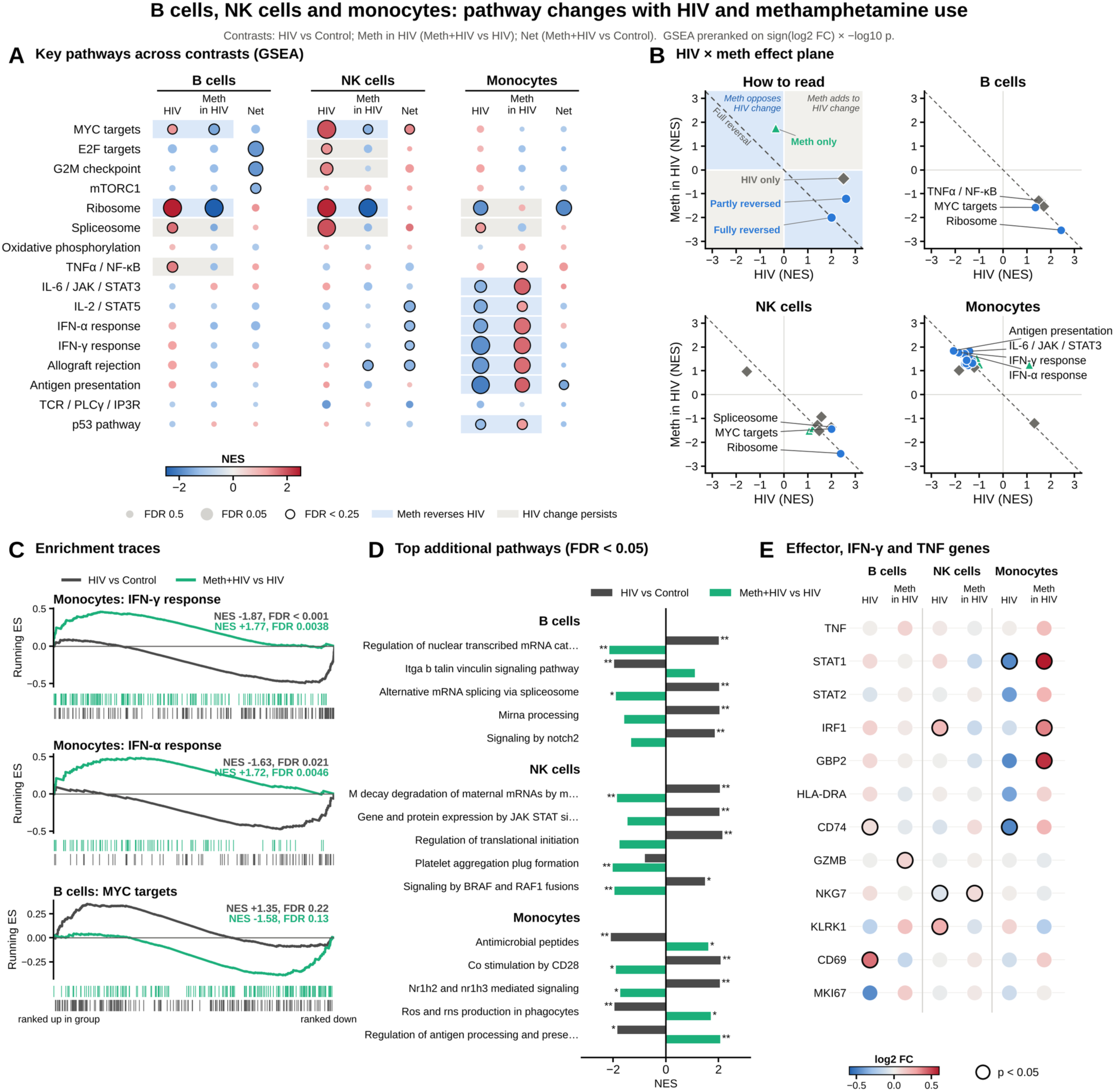
Methamphetamine use reverses HIV-associated suppression of interferon and antigen presentation programs in monocytes. GSEA of pseudobulk expression changes in B cells, NK cells and monocytes (n = 4 per group). Contrasts: HIV, HIV vs Control; Meth in HIV, Meth+HIV vs HIV; Net, Meth+HIV vs Control. **(A)** NES for 16 key Hallmark and KEGG pathways in each contrast. Color, NES (red, higher in the first-named group; blue, lower); size, −log10 FDR; black outline, FDR < 0.25; blue shading, reversed; gray shading, HIV change persists. **(B)** Effect plane relating the effects of HIV and of methamphetamine use within HIV on each pathway. Top left, key: x, NES in the HIV contrast; y, NES in the Meth in HIV contrast; blue quadrants, methamphetamine use moves a pathway in the opposite direction to HIV; gray quadrants, same direction; dashed line, full reversal (y = −x); example points show a fully reversed pathway (on the dashed line), a partly reversed pathway (between the dashed line and the x axis), a pathway changed with HIV only and a pathway changed with methamphetamine use only. Other panels, pathways changed at FDR < 0.25 in either contrast in B cells, NK cells and monocytes, plotted by NES in the HIV (x) and Meth in HIV (y) contrasts. Blue circles, reversed; green triangles, Meth in HIV only; gray diamonds, HIV only. Dashed line, full reversal (y = −x). **(C)** Running enrichment scores for monocyte IFN-γ response, monocyte IFN-α response and B cell MYC targets, with genes ranked from most increased (left) to most decreased (right), for the HIV (dark gray) and Meth in HIV (green) contrasts. Ticks mark pathway genes. **(D)** Additional pathways beyond the 16 shown in panel A. For each cell type, gene sets from Reactome, KEGG, GO Biological Process and PID with FDR < 0.05 in either contrast were ranked by FDR, and the top five that did not substantially overlap a panel A pathway or a set already shown are displayed (sets sharing more than half of their leading-edge genes were treated as overlapping). Bars, NES for the HIV (dark gray) and Meth in HIV (green) contrasts; * FDR < 0.25, ** FDR < 0.05. **(E)** Pseudobulk expression changes in the HIV and Meth in HIV contrasts for the 12 genes, from a predefined list of effector, interferon-stimulated and TNF pathway genes, with the smallest *p* value in either contrast across the three cell types. Color, log2 fold change; black outline, *p* < 0.05.

On the effect plane, B and NK cells and monocytes fell on opposite sides (**Fig. 3B**). In B and NK cells, HIV-associated changes lay in the lower right, and ribosome sat on the diagonal in both cell types (B cells, HIV NES = 2.43 and Meth in HIV NES = −2.52; NK cells, 2.37 and −2.47; all FDR < 0.001), so that ribosome programs no longer differed from controls in Meth+HIV participants. MYC targets were also reversed in both populations, although in NK cells the effect of methamphetamine use within HIV was smaller than the HIV effect and MYC targets remained enriched relative to controls. The remaining B and NK cell changes, including spliceosome and the NK cell cycle programs, were only significant in the HIV-only comparison. Monocytes showed the mirror image, with most points in the upper left. Of 19 altered monocyte pathways, 12 were reversed, including IFN-γ response, IFN-α response, allograft rejection, antigen processing and presentation, IL-6/JAK/STAT3 and IL-2/STAT5 signaling, and most of these lay close to the diagonal. Four pathways (inflammatory response, complement, DNA repair and TNFα signaling via NF-κB) changed only with methamphetamine use, all toward enrichment. Monocyte ribosome depletion was an HIV-only change and remained lower in Meth+HIV participants than in controls (NES = −1.94, FDR = 0.008).

Running enrichment scores show the monocyte interferon reversal for individual pathways (**Fig. 3C**). For both IFN-γ response and IFN-α response, the HIV trace dipped among the most down-regulated genes (NES = −1.87, FDR = 0.001 and NES = −1.63, FDR = 0.021), with *STAT1*, *CD74* and *IRF8* in the leading edge of the IFN-γ set, and the methamphetamine trace peaked among the most up-regulated genes to a similar degree (NES = 1.77, FDR = 0.004 and NES = 1.72, FDR = 0.005). As a result, neither interferon program differed from controls in Meth+HIV participants. For MYC targets in B cells, the two traces were smaller but likewise ran in opposite directions (HIV NES = 1.35, FDR = 0.22; Meth in HIV NES = −1.58, FDR = 0.13).

The additional gene sets again separated lymphocytes from monocytes (**Fig. 3D**). In B cells, HIV was associated with enrichment of mRNA decay, alternative splicing and miRNA processing gene sets, and deadenylation-dependent mRNA decay was reversed by methamphetamine use within HIV (HIV NES = 2.01; Meth in HIV NES = −2.14). NK cells showed HIV-associated enrichment of translation initiation, mRNA decay and IL-12-induced JAK-STAT gene expression sets, whereas methamphetamine use depleted platelet aggregation and RAF signaling sets. In monocytes, HIV was associated with depletion of antimicrobial peptide and phagocyte reactive oxygen and nitrogen species production sets and with enrichment of CD28 costimulation and LXR (*NR1H2* and *NR1H3*) signaling sets, and methamphetamine use increased regulation of antigen processing and presentation (NES = 2.06, FDR = 0.007). At the gene level, monocytes from PWH expressed less *STAT1* and less of the MHC class II chaperone *CD74*, and monocytes from Meth+HIV participants expressed more *STAT1*, *GBP2* and *IRF1* than those from HIV participants (**Fig. 3E**). In NK cells, HIV was associated with higher *KLRK1* and *IRF1* expression, and B cells from PWH expressed more of the activation marker *CD69*.

### Cytokine and transcription factor signatures in T cells point to opposing signals from HIV and methamphetamine use

GSEA shows which programs change but not what drives them. Because T cells showed the most varied relationships between the effects of HIV and of methamphetamine use within HIV (**Fig. 2**), we focused on T cells to identify the upstream signals that might underlie these programs. We scored cytokine response signatures (CytoSig^51^), signaling pathway footprints (PROGENy^52^) and transcription factor regulons (DoRothEA^53^), keeping only cytokines whose receptors were expressed by each cell type. Cytokine responses showed a consistent opposition between the two exposures. Of 36 cytokine responses that changed significantly in either contrast across the three T cell populations, 34 moved in opposite directions with HIV and methamphetamine use (**Fig. 4A**). The largest of these was the type III IFN response, which was weaker with HIV in CD4^+^ and CD8^+^ T cells (t = −8.1 and −7.9) and stronger with methamphetamine use (t = 6.9 and 4.9), with similar reversals of the IL-4, IL-10 and M-CSF responses. The TRAIL response showed the opposite pattern, rising with HIV in all three populations and falling with methamphetamine use. In CD4^+^ and γδ T cells, HIV was also associated with stronger TNF, IL-6 and IL-12 responses, which remained elevated relative to controls in Meth+HIV participants, and methamphetamine use raised the IL-2 response. CD8^+^ T cells showed the clearest reversal of interferon and IL-2 signaling. HIV was associated with weaker IFN-γ, type I IFN and IL-2 responses (t = −4.3, −2.5 and −6.8), and methamphetamine use strengthened the IFN-γ and IL-2 responses (t = 3.2 and 7.2).

**Figure 4.**
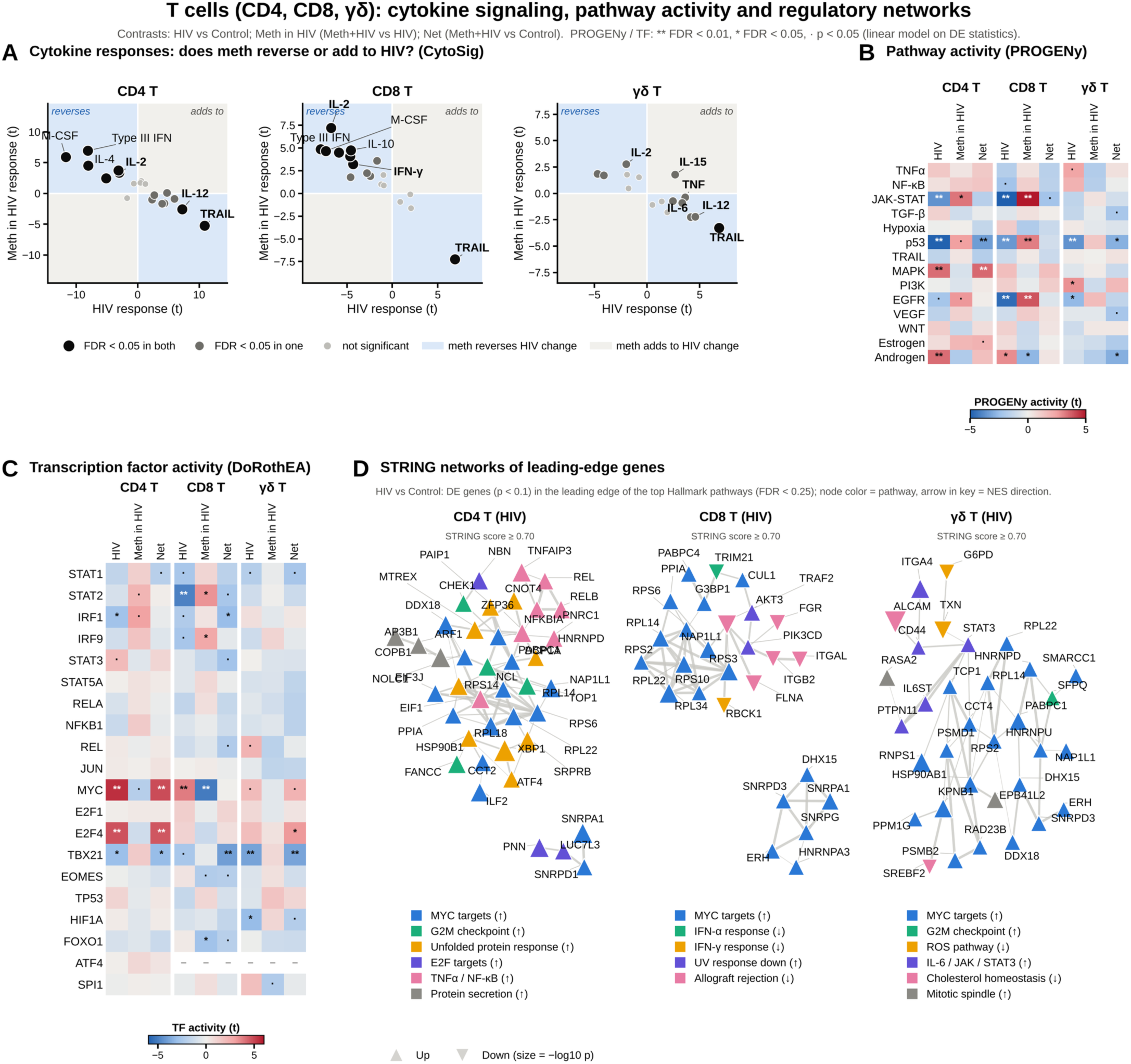
Cytokine response, signaling pathway and transcription factor activity in T cells. Activities were inferred from the gene-level statistic, sign(log2 fold change) × −log10 p, in CD4, CD8^+^ and γδ T cells (n = 4 per group). **(A)** CytoSig cytokine response scores (t statistic) in the HIV (x) and Meth in HIV (y) contrasts, for cytokines whose receptor genes were detected (CD4^+^ T, 19 of 24; CD8^+^ T, 18; γδ T, 16). Positive values, stronger response in the first-named group. Blue quadrants, methamphetamine use moves the response opposite to HIV; gray quadrants, same direction. Black, FDR < 0.05 in both contrasts; dark gray, in one; light gray, neither. **(B)** PROGENy activity for 14 signaling pathways in the HIV, Meth in HIV and Net contrasts. **(C)** DoRothEA activity for 20 immune and growth-related transcription factors (regulons A to C); dash, fewer than 10 measured targets. In B and C, red, higher activity in the first-named group; blue, lower; ** FDR < 0.01, * FDR < 0.05, · *p* < 0.05. **(D)** STRING networks for the HIV contrast, built from leading-edge genes of the top Hallmark pathways (FDR < 0.25; gene *p* < 0.1). Node color, pathway; up and down triangles, increased and decreased genes; node size, −log10 p; edge width, STRING score (threshold shown in each panel).

Pathway and transcription factor activities showed similar opposing patterns (**Fig. 4B,C**). In CD8^+^ T cells, HIV was associated with lower JAK-STAT (t = −6.4), EGFR and p53 pathway activity and lower STAT2 activity (t = −4.9), and methamphetamine use reversed each of these (JAK-STAT, t = 5.1; STAT2, t = 3.3). MYC activity, which was higher with HIV in CD8^+^ T cells (t = 3.8), fell with methamphetamine use (t = −5.1), matching the reversal of MYC targets by GSEA. In CD4^+^ T cells, HIV was associated with higher MYC, E2F4 and MAPK activity and lower JAK-STAT and p53 activity. Methamphetamine use partially restored JAK-STAT activity, but MYC, E2F4 and MAPK activity remained higher than in controls (Net t = 4.8, 4.6 and 3.5). T-bet (TBX21) activity was lower with HIV in CD4^+^ and γδ T cells (t = −3.0 and −3.9) and was lower in Meth+HIV participants than in controls in all three T cell populations (Net t = −3.1, −4.0 and −3.8).

To see how the genes behind these HIV-associated programs relate to one another, we built STRING interaction networks from the leading-edge genes of the top Hallmark pathways in each T cell population (**Fig. 4D**). In CD4^+^ T cells, a large MYC target and translation cluster, which included E2F target, G2M checkpoint and unfolded protein response genes, connected to a separate NF-κB cluster of increased genes (*NFKBIA*, *REL*, *RELB*, *TNFAIP3* and *ZFP36*). In CD8^+^ T cells, increased ribosomal and MYC target genes linked to decreased allograft rejection and interferon response genes, including *ITGB2*, *ITGAL*, *FGR*, *TRAF2* and *G3BP1*, with a separate spliceosome module. The γδ T cell network centered on *STAT3*, connecting the IL-6/JAK/STAT3 genes *IL6ST*, *CD44* and *PTPN11* to a MYC target and translation module. The same analyses for B cells, NK cells and monocytes are shown in Supplementary Fig. S4, and all activity scores and network genes are provided in the supplementary tables.

### Central memory CD8^+^ T cells of Meth+HIV participants lose type 1 effector programs

Because CD8^+^ T cells are central to HIV-associated neuroinflammation, we next asked which CD8^+^ T cell subsets carried these changes, separating naive, central memory, GZMK⁺ memory, CX3CR1⁺ effector memory and cytotoxic effector cells (**Fig. 5**). The most notable change was in central memory cells. Compared with controls, central memory CD8^+^ T cells from Meth+HIV participants had weaker IFN-γ response, complement, allograft rejection and cytotoxicity programs, weaker IFN-γ and TNF response signatures, and lower activity of T-bet and EOMES, the transcription factors that drive type 1 effector function (**Fig. 5A-C**). Part of this loss was already present with HIV, and methamphetamine use deepened it, further lowering T-bet activity and expression of the effector genes *GNLY*, *CX3CR1*, *PRDM1* and *ZEB2* (**Fig. 5C,D**). Memory subsets showed a different change, with HIV-associated MYC target programs persisting in GZMK⁺ and CX3CR1⁺ memory cells of Meth+HIV participants (**Fig. 5A,B**). In every CD8^+^ T cell subtype, the IL-2 response fell with HIV and rose with methamphetamine use, as it did in CD8^+^ T cells overall (**Fig. 5C**).

**Figure 5.**
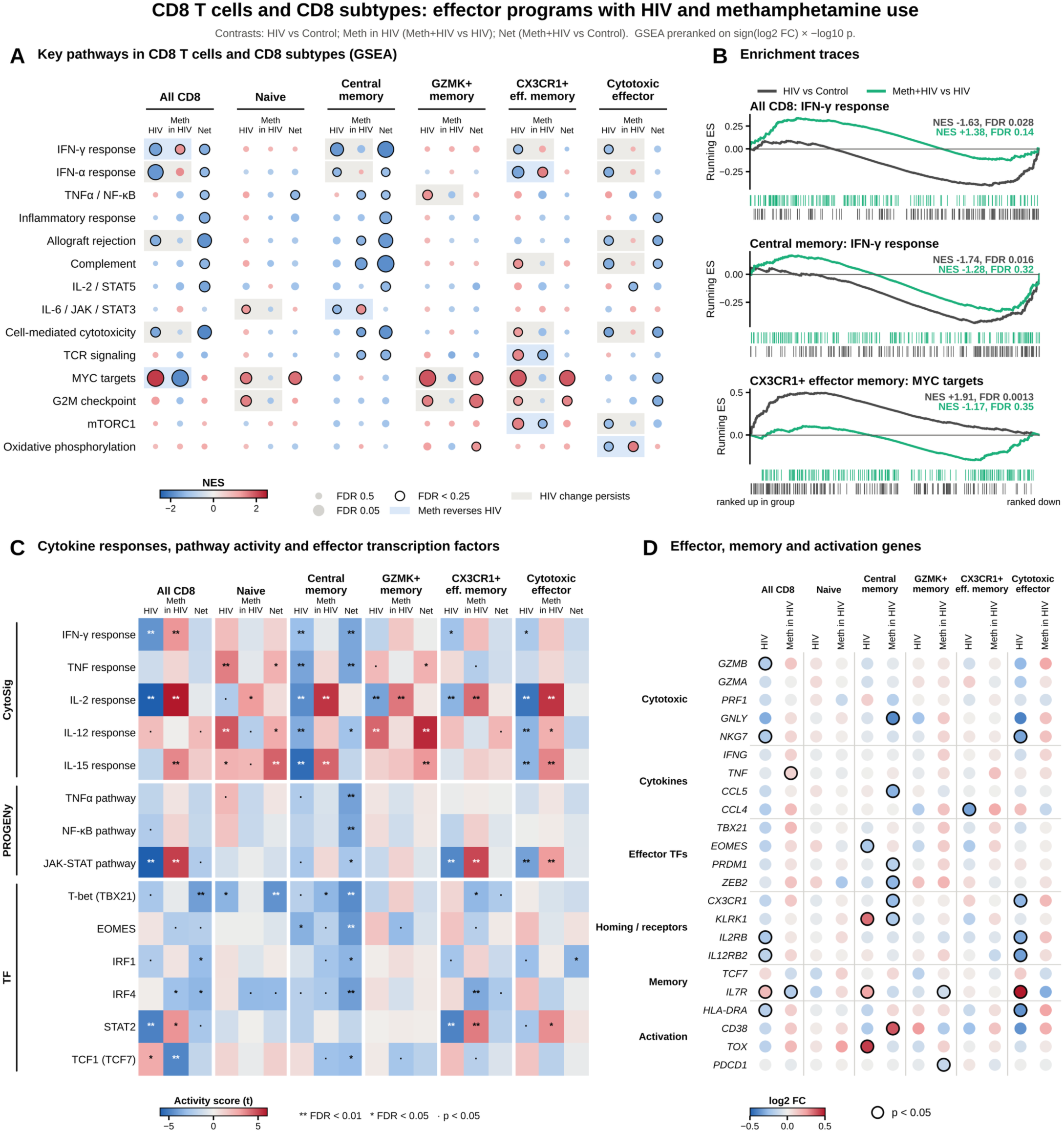
Central memory CD8^+^ T cells of Meth+HIV participants lose type 1 effector programs. Pseudobulk analyses of all CD8^+^ T cells and five CD8^+^ T cell subtypes (naive, central memory, GZMK⁺ memory, CX3CR1⁺ effector memory and cytotoxic effector; n = 4 per group). Contrasts: HIV, HIV vs Control; Meth in HIV, Meth+HIV vs HIV; Net, Meth+HIV vs Control. **(A)** GSEA NES for 14 Hallmark and KEGG pathways. Color, NES (red, higher in the first-named group; blue, lower); size, −log10 FDR; black outline, FDR < 0.25; blue shading, reversed; gray shading, HIV change persists. Cell-mediated cytotoxicity is the KEGG natural killer cell-mediated cytotoxicity set. **(B)** Running enrichment scores for IFN-γ response in all CD8^+^ T cells, IFN-γ response in central memory cells and MYC targets in CX3CR1⁺ effector memory cells, for the HIV (dark gray) and Meth in HIV (green) contrasts. **(C)** Activity scores (t statistic) for selected cytokine responses (CytoSig), signaling pathways (PROGENy) and transcription factors (DoRothEA). Red, higher in the first-named group; blue, lower; ** FDR < 0.01, * FDR < 0.05, · *p* < 0.05. **(D)** Pseudobulk expression changes in the HIV and Meth in HIV contrasts for a predefined panel of cytotoxic, cytokine, effector transcription factor, homing and receptor, memory and activation genes, grouped by function. Color, log2 fold change; black outline, *p* < 0.05.

### Monocyte antigen presentation programs suppressed in HIV rise with methamphetamine use and parallel interferon signatures in CD8^+^ T cells

Monocytes showed the clearest reversal of any population, and the reversed programs centered on interferon response and antigen presentation (**Fig. 3**). To ask how far this extended, we examined antigen presentation gene sets across the GO, KEGG and Reactome collections (**Fig. 6A**). With HIV, monocytes showed lower enrichment of MHC class II and class I presentation, peptide loading, lysosomal processing and IFN-γ signaling gene sets, including KEGG antigen processing and presentation (NES = −2.08, FDR = 0.002), MHC protein complex (NES = −1.91, FDR = 0.033) and lysosome (NES = −1.75, FDR = 0.031).

**Figure 6.**
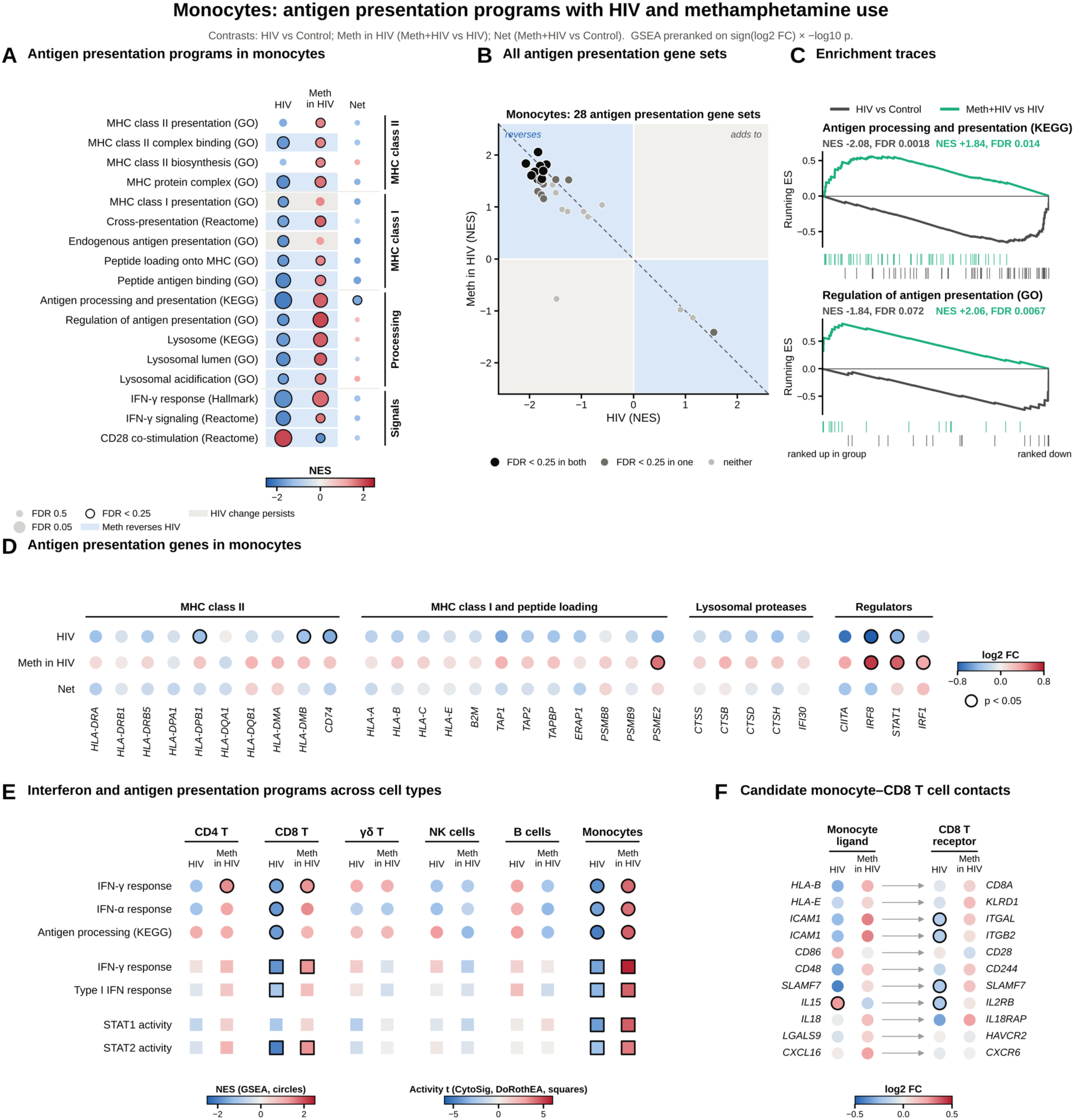
Monocyte antigen presentation programs suppressed in HIV rise with methamphetamine use and parallel interferon signatures in CD8^+^ T cells. Pseudobulk analyses of monocytes and, in E and F, other PBMC populations (n = 4 per group). Contrasts: HIV, HIV vs Control; Meth in HIV, Meth+HIV vs HIV; Net, Meth+HIV vs Control. **(A)** GSEA NES in monocytes for 17 gene sets covering MHC class II presentation, MHC class I presentation and peptide loading, antigen processing and lysosomal function, and the IFN-γ and CD28 co-stimulatory signals linked to presentation, from the GO, KEGG, Reactome and Hallmark collections. Color, NES (red, higher in the first-named group; blue, lower); size, −log10 FDR; black outline, FDR < 0.25; blue shading, reversed; gray shading, HIV change persists. **(B)** All 28 antigen presentation gene sets in the GO, KEGG, Reactome, BioCarta and PID collections (names containing antigen processing, antigen presentation, MHC, peptide antigen or cross-presentation), plotted by NES in the HIV (x) and Meth in HIV (y) contrasts in monocytes. Blue quadrants, methamphetamine use moves the gene set in the opposite direction to HIV; gray quadrants, same direction; dashed line, full reversal (y = −x). Black, FDR < 0.25 in both contrasts; dark gray, in one; light gray, neither. **(C)** Running enrichment scores in monocytes for KEGG antigen processing and presentation and GO regulation of antigen processing and presentation, with genes ranked from most increased (left) to most decreased (right), for the HIV (dark gray) and Meth in HIV (green) contrasts. Ticks mark gene set genes. **(D)** Pseudobulk log2 fold changes in monocytes for MHC class II, MHC class I and peptide loading, lysosomal protease and transcriptional regulator genes in the HIV, Meth in HIV and Net contrasts. Color, log2 fold change; black outline, *p* < 0.05. **(E)** Interferon and antigen presentation programs in each PBMC population in the HIV and Meth in HIV contrasts. Circles, GSEA NES for Hallmark IFN-γ response, Hallmark IFN-α response and KEGG antigen processing and presentation; squares, CytoSig IFN-γ and type I IFN response scores and DoRothEA STAT1 and STAT2 activity (t statistic). Color, NES or t (red, higher in the first-named group; blue, lower); black outline, FDR < 0.25 (GSEA) or FDR < 0.05 (activity scores). **(F)** Candidate contacts between monocytes and CD8^+^ T cells: canonical ligand-receptor pairs for antigen presentation, adhesion, co-stimulation, SLAM family, cytokine, inhibitory and chemokine signaling, with the ligand detected in monocytes and the receptor in CD8^+^ T cells. Left, log2 fold change of the ligand in monocytes; right, log2 fold change of the receptor in CD8^+^ T cells, in the HIV and Meth in HIV contrasts. Black outline, *p* < 0.05.

Methamphetamine use shifted each of these in the opposite direction (antigen processing and presentation, NES = 1.84, FDR = 0.014; lysosome, NES = 1.87, FDR = 0.013), and regulation of antigen processing and presentation was strongly enriched with methamphetamine use (NES = 2.06, FDR = 0.007). In Meth+HIV participants, only KEGG antigen processing and presentation remained marginally lower than in controls (Net NES = −1.57, FDR = 0.24). CD28 co-stimulation showed the opposite pattern, increasing with HIV (NES = 2.08, FDR = 0.001) and decreasing with methamphetamine use (NES = −1.90, FDR = 0.23). The shift was not limited to these selected gene sets. Of all 28 antigen presentation gene sets, 25 were lower with HIV (15 at FDR < 0.25) and 24 were higher with methamphetamine use (12 at FDR < 0.25), and 24 fell in the reversal quadrants of the effect plane (**Fig. 6B**). Running enrichment scores showed the HIV and methamphetamine contrasts acting on the same genes in opposite directions (**Fig. 6C**).

At the level of individual genes, changes were modest but consistent across the antigen presentation machinery (**Fig. 6D**). Among 27 genes related to MHC class II presentation, MHC class I presentation and peptide loading, and lysosomal protease activity, 26 were lower in monocytes from PWH than in controls, whereas 25 were higher in Meth+HIV than in HIV participants. Significant HIV-associated decreases (*p* < 0.05) were in *CD74*, *HLA-DPB1* and *HLA-DMB*. Transcriptional regulators of antigen presentation followed the same pattern. *IRF8* and *STAT1* were lower with HIV (log2 fold change = −1.06 and −0.47) and higher with methamphetamine use (0.70 and 0.59), *IRF1* was higher with methamphetamine use, and the MHC class II transactivator *CIITA* tended to be lower with HIV (log2 fold change = −0.72, *p* < 0.1).

Antigen presentation by monocytes shapes CD8^+^ T cell activation, and IFN-γ from activated CD8^+^ T cells in turn induces MHC expression in monocytes. We therefore compared these programs across cell types (**Fig. 6E**). Monocytes and CD8^+^ T cells were the only populations in which HIV was associated with lower IFN-γ response, IFN-α response and antigen processing programs, together with weaker IFN-γ and type I IFN response signatures and lower STAT2 activity. For example, the IFN-γ response signature was weaker with HIV in monocytes (t = −3.8) and CD8^+^ T cells (t = −4.3) and stronger with methamphetamine use (t = 5.7 and 3.2). Methamphetamine use reversed these changes fully in monocytes and partly in CD8^+^ T cells, where the IFN-γ response and STAT2 activity rose but antigen processing programs did not change significantly. Across the 28 antigen presentation gene sets, CD8^+^ T cells also showed HIV-associated decreases in 26 (11 at FDR < 0.25), whereas none was significantly lower with HIV in CD4^+^ T, γδ T, NK or B cells. Among candidate contact molecules (**Fig. 6F**), the LFA-1 chains *ITGAL* and *ITGB2* and the SLAM family receptor *SLAMF7* were lower in CD8^+^ T cells from PWH (*p* < 0.05), and their monocyte partners *ICAM1* and *SLAMF7* were also lower with HIV, although not significantly. Together, these data point to coordinated changes in monocyte antigen presentation and CD8^+^ T cell interferon signaling with HIV and methamphetamine use.

### Plasma from PWH shows cytotoxic lymphocyte activation that rises further with methamphetamine use

We next asked whether these cellular changes were reflected in circulating proteins, using the NULISAseq Inflammation panel in a larger set of participants (**Fig. 7**). HIV was associated only with increases in this panel (22 proteins higher at *p* < 0.05 and none lower; **Fig. 7A**), and the strongest were markers of cytotoxic and type 1 lymphocyte activation. The NKG2D receptor KLRK1 was the only protein to reach FDR < 0.05 (Δ NPQ = 0.74, FDR = 0.025), and granzyme B (Δ NPQ = 1.14), IFN-γ (0.94), granzyme A, CXCL9, TNF and the receptors IL12RB1 and IL18R1 were also higher in PWH than in controls (all *p* < 0.05). PWH also had higher levels of the T cell activation and checkpoint proteins IL2RA (CD25), CTLA4, LAG3 and PD-1 and of the B cell survival receptor BCMA (TNFRSF17).

**Figure 7.**
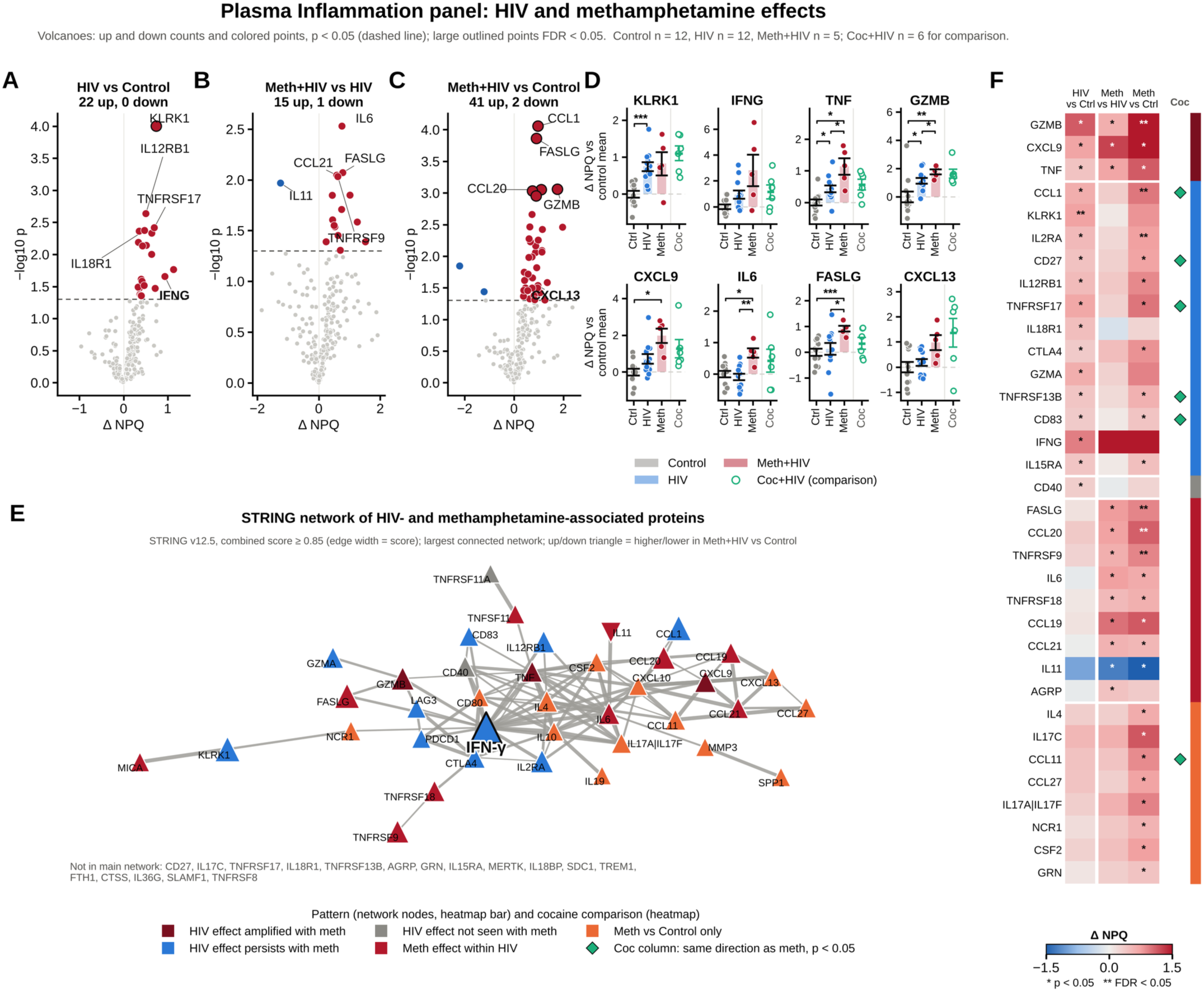
Plasma inflammation proteins with HIV and methamphetamine use. NULISAseq Inflammation panel (249 proteins; NPQ, log2 scale) in Control (n = 12), HIV (n = 12) and Meth+HIV (n = 5) participants, with Coc+HIV (n = 6) shown for comparison. Groups were compared with Welch t tests; Δ NPQ, difference in group means. **(A to C)** Volcano plots of HIV vs Control **(A)**, Meth+HIV vs HIV **(B)** and Meth+HIV vs Control **(C)**. Numbers above each plot, proteins increased and decreased at *p* < 0.05; red and blue points, increased and decreased at *p* < 0.05; outlined points, FDR < 0.05; dashed line, *p* = 0.05. **(D)** KLRK1, IFN-γ, TNF, granzyme B, CXCL9, IL-6, FASLG and CXCL13 as Δ NPQ relative to the control mean (mean ± SEM; points, individual participants). Coc+HIV is shown at right as open green points with the mean (green line) ± SEM. Brackets, pairwise Welch t tests among Control, HIV and Meth+HIV with Holm-Šídák adjustment: * *p* < 0.05, ** *p* < 0.01, *** *p* < 0.001, **** *p* < 0.0001. **(E)** STRING network of proteins with a pattern of change in F. Node color, pattern; node size, smallest *p* value across comparisons; up and down triangles, higher and lower in Meth+HIV than Control; IFN-γ, the most connected protein, is drawn larger with a black outline and bold label; proteins outside the main network are listed below. **(F)** Δ NPQ for proteins with a pattern of change in each comparison (* *p* < 0.05; ** FDR < 0.05). Green diamonds in the Coc column mark proteins also changed by cocaine use in the same direction (*p* < 0.05). Side bar, pattern: amplified (HIV vs Control and Meth+HIV vs HIV both *p* < 0.05, same direction); persists (HIV vs Control *p* < 0.05, Meth+HIV vs HIV p ≥ 0.05 and Meth+HIV vs Control *p* < 0.10, same direction); not seen with methamphetamine (other HIV vs Control changes); methamphetamine within HIV only; and Meth+HIV vs Control only.

Methamphetamine use did not reverse any of these HIV-associated changes. Of the 22 proteins increased with HIV, granzyme B, CXCL9 and TNF rose further with methamphetamine use, 17 remained higher in Meth+HIV participants than in controls and only 2 were no longer elevated (**Fig. 7B, F**). IFN-γ was higher still in Meth+HIV participants, although this increase was variable and did not reach significance (**Fig. 7D**). Methamphetamine use also brought changes that HIV alone had not (15 proteins higher and 1 lower in Meth+HIV than HIV participants), most prominently IL-6 (Δ NPQ = 0.76, *p* = 0.003), FASLG, the co-stimulatory receptors 4-1BB (TNFRSF9) and GITR (TNFRSF18), the lymphoid chemokines CCL19, CCL20 and CCL21 and the NKG2D ligand MICA, whereas IL-11 was lower (**Fig. 7B**). Pairwise comparisons showed a stepwise rise from Control to HIV to Meth+HIV for TNF and granzyme B, whereas IL-6 and FASLG were higher in Meth+HIV participants than in either other group (**Fig. 7D**). Taken together, Meth+HIV participants differed from controls in 43 proteins (41 higher and 2 lower), six of which reached FDR < 0.05 (CCL1, FASLG, granzyme B, IL2RA, 4-1BB and CCL20), and they also had higher CXCL13 (Δ NPQ = 0.97, *p* = 0.029; **Fig. 7C, D**).

These proteins formed a single network in which a cytotoxic and co-stimulatory module built around IFN-γ, TNF and IL-6 connected to a lymphoid chemokine module containing CXCL9 and CXCL13 (**Fig. 7E**). IFN-γ was the most connected protein in this network, linked directly to 18 of the 37 other proteins, including TNF, IL-6, granzyme B, FASLG, CXCL9, CXCL10 and the checkpoint receptors CTLA4, LAG3 and PD-1.

Cocaine use moved 15 of the 16 proteins changed by methamphetamine use within HIV in the same direction, but none of these shifts reached *p* < 0.05 in Coc+HIV compared with HIV participants, and across the panel the methamphetamine and cocaine effects were only weakly correlated (r = 0.12). Relative to controls, however, Coc+HIV participants differed in 33 proteins, and 9 proteins that changed in Meth+HIV participants relative to controls, including CCL1, CD27, BCMA and CCL11, changed in the same direction with cocaine use (**Fig. 7F**).

### Methamphetamine use is associated with higher plasma neurofilament heavy chain

The CNS panel gave a different picture (**Fig. 8**). HIV alone changed only five proteins, raising TNF, the neuronal adhesion protein contactin-2 (CNTN2) and IGF1R and lowering CCL2 and IL-15 (**Fig. 8A**), whereas methamphetamine use within HIV changed 13 (11 higher and 2 lower; **Fig. 8B**). The largest change in either plasma panel was a rise in neurofilament heavy chain (NEFH), a structural protein released from injured axons (Δ NPQ = 4.48 in Meth+HIV compared with HIV participants, FDR = 0.004). Methamphetamine use also increased the axon guidance protein SLIT2 (FDR = 0.042), the amyloid-β peptides Aβ40 and Aβ42 (Δ NPQ = 0.59 and 0.37; *p* = 0.007 and 0.008), the kallikrein KLK6, IGFBP7, TREM1 and the cytokines TNF, IL-6, IL-2 and GM-CSF, and lowered SFRP1 and SOD1.

**Figure 8.**
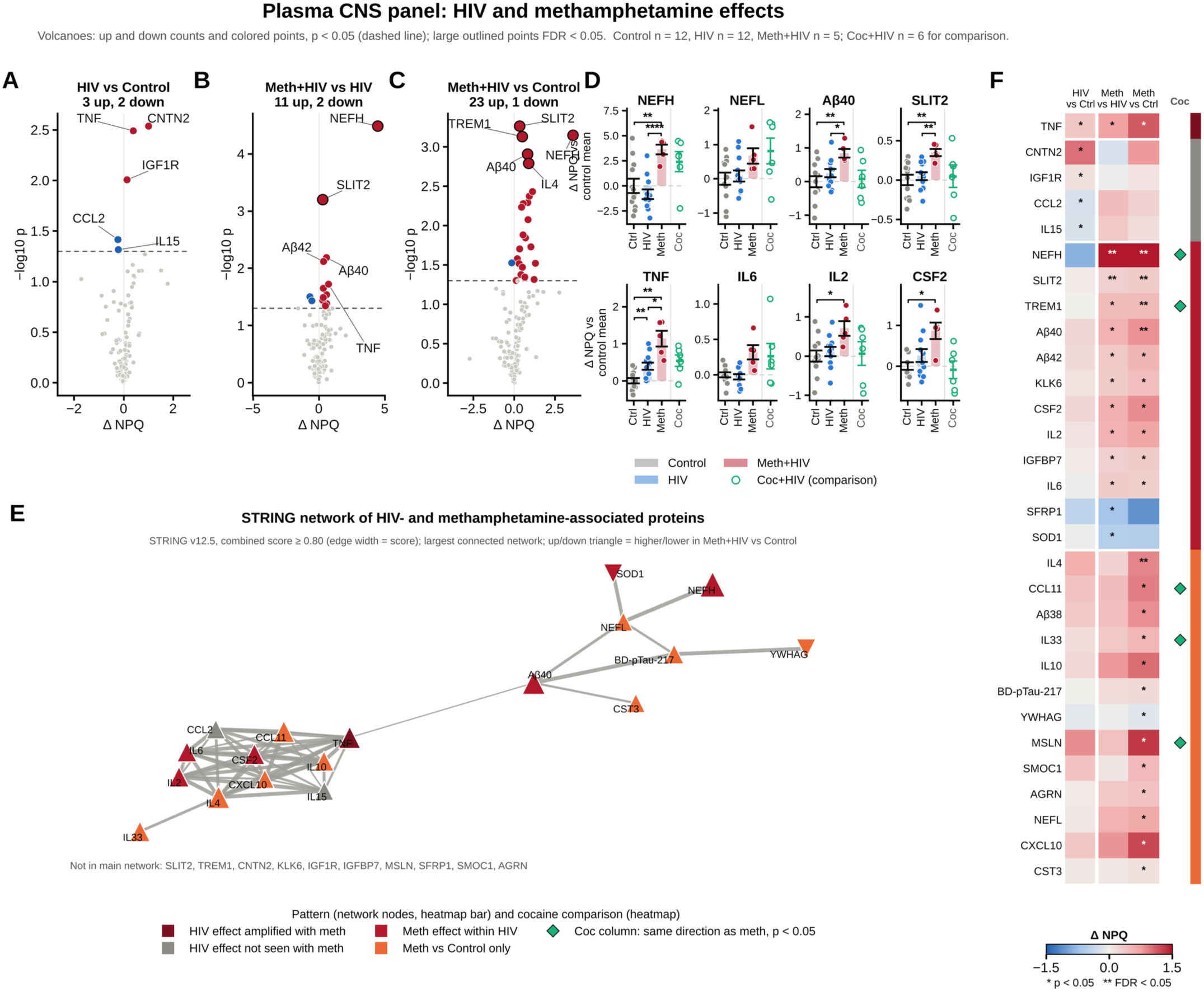
Plasma CNS proteins with HIV and methamphetamine use. NULISAseq CNS panel (131 proteins; NPQ, log2 scale) in Control (n = 12), HIV (n = 12) and Meth+HIV (n = 5) participants, with Coc+HIV (n = 6) shown for comparison. Groups were compared with Welch t tests; Δ NPQ, difference in group means. **(A to C)** Volcano plots of HIV vs Control **(A)**, Meth+HIV vs HIV **(B)** and Meth+HIV vs Control **(C)**. Numbers above each plot, proteins increased and decreased at *p* < 0.05; red and blue points, increased and decreased at *p* < 0.05; outlined points, FDR < 0.05; dashed line, *p* = 0.05. **(D)** NEFH, NEFL, Aβ40, SLIT2, TNF, IL-6, IL-2 and GM-CSF as Δ NPQ relative to the control mean (mean ± SEM; points, individual participants). Coc+HIV is shown at right as open green points with the mean (green line) ± SEM. Brackets, pairwise Welch t tests among Control, HIV and Meth+HIV with Holm-Šídák adjustment: * *p* < 0.05, ** *p* < 0.01, *** *p* < 0.001, **** *p* < 0.0001. **(E)** STRING network of proteins with a pattern of change in F (Aβ and pTau species mapped to APP and MAPT). Node color, pattern; node size, smallest *p* value across comparisons; up and down triangles, higher and lower in Meth+HIV than Control; proteins outside the main network are listed below. **(F)** Δ NPQ for proteins with a pattern of change in each comparison (* *p* < 0.05; ** FDR < 0.05). Green diamonds in the Coc column mark proteins also changed by cocaine use in the same direction (*p* < 0.05). Side bar, pattern: amplified (HIV vs Control and Meth+HIV vs HIV both *p* < 0.05, same direction); not seen with methamphetamine (HIV vs Control *p* < 0.05 without a same-direction Meth+HIV vs Control change at *p* < 0.10); methamphetamine within HIV only; and Meth+HIV vs Control only.

Amyloid-β peptides did not differ significantly between HIV and Control participants (Aβ40, Δ NPQ = 0.25, *p* = 0.23; Aβ42, Δ NPQ = 0.21, *p* = 0.25), so their increase was associated with methamphetamine use rather than with HIV alone, and Aβ38 was also higher in Meth+HIV participants than in controls (*p* = 0.005). Compared with controls, Meth+HIV participants differed in 24 CNS panel proteins, including higher NEFH, NEFL, SLIT2, Aβ40 and TREM1 (**Fig. 8C**). NEFH, Aβ40 and SLIT2 were higher in Meth+HIV participants than in either other group, whereas TNF rose step by step from Control to HIV to Meth+HIV (**Fig. 8D**). In the interaction network, neurofilaments, Aβ40 and pTau-217 formed a neuronal module that connected through Aβ40 to a cytokine module of TNF, IL-6, IL-2 and GM-CSF (**Fig. 8E**).

Cocaine use also raised NEFH (Δ NPQ = 3.25 in Coc+HIV compared with HIV participants, *p* = 0.021) and TREM1, whereas the increases in SLIT2, amyloid-β, IL-2 and GM-CSF were not seen with cocaine use (**Fig. 8D, F**). Across the CNS panel, methamphetamine and cocaine effects were moderately correlated (r = 0.31), and 6 of the 13 methamphetamine-associated proteins changed in the same direction with cocaine use.

## Discussion

HIV-associated cognitive impairment reflects interacting viral, inflammatory, glial, and systemic processes^63^, and stimulant use adds to this burden. In this study, we combined single-cell transcriptomics of PBMCs with targeted plasma proteomics to define how methamphetamine use shapes immune dysregulation in PWH. Three main findings emerge. First, methamphetamine use acts in a lineage-specific manner. HIV-associated proliferative and TNF signaling programs persist in CD4^+^ and γδ T cells of Meth+HIV participants, whereas in monocytes and CD8^+^ T cells methamphetamine use shifts HIV-associated programs in the opposite direction, most clearly interferon and antigen presentation programs in monocytes and IFN-γ, IL-2 and MYC programs in CD8^+^ T cells. Within the CD8^+^ T cell compartment, central memory cells of Meth+HIV participants show reduced type 1 effector programming. Second, the monocyte shift spans the antigen presentation machinery, from MHC class II and class I presentation to peptide loading, lysosomal processing and the regulators IRF8 and STAT1, and parallels interferon signatures in CD8^+^ T cells. Third, plasma from PWH carries a cytotoxic lymphocyte activation signature that is further elevated with methamphetamine use, together with increases in IL-6, CXCL13 and neurofilament heavy chain.

The persistence of T cell activation programs is consistent with the chronic immune activation that characterizes treated HIV infection^12,13^. Single-cell studies of PBMCs have shown that ART restores much of the HIV-perturbed immune transcriptome but that some changes persist^64^, and that T cell dysfunction persists in PWH with incomplete immune recovery^65^. In CD4^+^ and γδ T cells, HIV was associated with enrichment of MYC, E2F and G2M programs, strong TNF, IL-12 and IL-6 response signatures and NF-κB activation, together with reduced expression of proximal TCR signaling genes. Reduced *LCK*, *ZAP70* and *LCP2* alongside heightened NF-κB and cytokine-driven programs suggests a shift from antigen receptor-driven toward bystander, cytokine-driven activation. Methamphetamine use did not normalize these programs and instead increased IL-2 response signatures across lymphocyte populations, in line with reports that methamphetamine alters T cell function in PWH on ART^24^ and impairs T cell subset progression and polyfunctionality in chronic viral infection^22,23^.

The CD8^+^ T cell compartment showed two distinct effects. In CD8^+^ T cells as a whole, methamphetamine use opposed HIV-associated suppression of IFN-γ, IL-2 and JAK-STAT signatures and HIV-associated enrichment of MYC and translational programs. Within the compartment, however, central memory cells of Meth+HIV participants showed reduced IFN-γ and TNF response programs, reduced T-bet and EOMES activity, and lower expression of the effector differentiation genes PRDM1, ZEB2, CX3CR1 and GNLY. T-bet and EOMES drive the effector and memory differentiation of CD8^+^ T cells, and central memory cells provide the pool from which effector cells are regenerated^66,67^. A loss of type 1 effector programming in this pool is in line with reports that methamphetamine impairs CD8^+^ T cell subset progression and polyfunctionality in chronic viral infection^22,23^. In parallel, HIV-associated MYC programs persisted in GZMK+ and CX3CR1+ memory CD8^+^ T cells of Meth+HIV participants, consistent with the durable expansion of CD8^+^ T cells in PWH^14–16^ and with methamphetamine-associated CD8^+^ T cell proliferation across memory subsets^24^. Cytolytic CD8^+^ T cells contribute to control of HIV replication in lymphoid tissue^68^, whereas IFN-γ-producing, non-cytolytic CD8^+^ T cells in CSF are linked to neurocognitive impairment in PWH^21^, and ongoing T cell activation in CSF tracks cognitive decline^20^. Lower interferon response signatures in circulating CD8^+^ T cells from PWH contrasted with higher plasma IFN-γ, which may reflect IFN-γ production at sites other than the circulating compartment or reduced sensitivity of circulating cells to chronic interferon exposure. Because the CD8^+^ T cells most closely linked to neurocognitive impairment in PWH are those producing IFN-γ in CSF^21^, this dissociation raises the possibility that the IFN-γ signal relevant to the CNS is compartmentalized rather than systemic. Paired blood and CSF profiling in the same participants, combined with measures of brain circuit function, will be needed to resolve where this activity resides and how the rebalancing of CD8^+^ T cell programs we observed in blood relates to CD8^+^ T cells in the CNS.

In monocytes, HIV was associated with suppression of interferon response, antigen presentation and JAK-STAT signaling, and methamphetamine use shifted each of these programs in the opposite direction, with concordant evidence from GSEA, cytokine response signatures, pathway footprints, and transcription factor activity. Interferon-stimulated programs are prominent in the earliest systemic immune response to HIV^69^, and type I interferon signaling persists in astrocytes, microglia and endothelial cells of the brain in PWH with cognitive impairment despite ART^70^. The monocyte programs shifted by methamphetamine use within HIV included both type I and IFN-γ (type II) responses. This pattern should not be read as restoration of healthy monocyte function. Methamphetamine use also enriched IL-6/JAK/STAT3 and inflammatory response programs and raised TNF response signatures, and the monocyte network for the methamphetamine contrast linked STAT1 to the Toll-like receptor genes *TLR1*, *TLR2* and *TLR6*, *MYD88* and *NLRP3*. These features are consistent with methamphetamine-induced innate immune activation superimposed on HIV-associated changes, in keeping with increased Toll-like receptor signaling and inflammatory programs reported in SIV infection and in brain myeloid cells of SIV-infected macaques exposed to methamphetamine^26–28^. Interferon and antigen presentation programs in myeloid cells are central to HIV neuropathogenesis^31,32,34,37^, so methamphetamine-driven increases in these pathways could promote myeloid activation in the CNS, particularly given methamphetamine-induced disruption of the blood-brain barrier^29,30^. Monocytes and the macrophages and dendritic cells derived from them present antigen to CD8^+^ T cells through MHC class I and cross-presentation and supply the co-stimulatory and adhesion signals that shape CD8^+^ T cell activation and effector differentiation. The breadth of the monocyte shift, which extended across MHC class II and class I presentation, peptide loading, lysosomal processing and the regulators IRF8 and STAT1, together with parallel changes in interferon signatures and in LFA-1 and SLAMF7 expression in CD8^+^ T cells, suggests that changes in monocyte antigen presentation may feed into the CD8^+^ T cell changes we observed. In this model, HIV-associated suppression of monocyte antigen presentation would limit CD8^+^ T cell stimulation and interferon signaling, and methamphetamine use would restore it. Functional studies of antigen presentation and CD8^+^ T cell activation in co-culture and in vivo will be needed to test this.

The plasma proteome extended these findings to soluble mediators. Plasma proteins can serve as accessible indicators of CNS-relevant immune activity in PWH^71^, and a previous plasma proteomic study found that proteins involved in complement, coagulation and oxidative stress changed over time in PWH with ongoing methamphetamine use^72^. Using a targeted platform with high sensitivity for low-abundance cytokines and immune receptors^59^, we found that HIV was associated with increases in KLRK1, IFN-γ, TNF, granzyme B, CXCL9, IL12RB1 and IL18R1, a profile of type 1 and cytotoxic lymphocyte activation consistent with persistent CD8^+^ T cell activation in treated HIV^14,16,17^. CXCL9 shares the receptor CXCR3 with CXCL10, which recruits activated CD8^+^ T cells into tissues and the CNS^73–76^. Methamphetamine use did not reverse any HIV-associated plasma changes; instead, TNF, granzyme B and CXCL9 were further increased, and IL-6, FASLG and 4-1BB were higher. Meth+HIV participants also had higher CXCL13 than controls. CXCL13 recruits CXCR5+ immune cells to sites of inflammation and can act on neural circuits^39–42^, and our recent work identified CXCL13-mediated CD8^+^ T cell recruitment as necessary for opioid self-administration^38^. The increase in plasma CXCL13 in PWH who use methamphetamine therefore nominates a candidate pathway linking stimulant use to CD8^+^ T cell recruitment in PWH. In the CNS panel, methamphetamine use was associated with a marked increase in NEFH, accompanied by increases in NEFL and Aβ40, suggesting ongoing neuroaxonal injury^77,78^. Plasma Aβ40 and Aβ42 were also higher with methamphetamine use, whereas HIV alone and cocaine use were not associated with a significant change. PWH are at increased risk of cerebral amyloid-β plaque deposition^79^, so a methamphetamine-associated rise in circulating amyloid-β may add to an amyloid-related burden in a population already at elevated risk of Alzheimer-type pathology. Most methamphetamine-associated plasma changes moved in the same direction with cocaine use, and NEFH was raised by both stimulants, whereas the increases in SLIT2, Aβ40 and IL-2 were specific to methamphetamine.

This study has several limitations. The design is cross-sectional, so causal relationships cannot be inferred. The cohort did not include people without HIV who use methamphetamine, so methamphetamine effects are defined within HIV and cannot be separated from interactions with HIV. Group sizes, particularly for the methamphetamine group in the proteomic analyses, were modest, meaning many of the results will need to be replicated with larger sample sizes to make these results more generalizable. PBMC profiling captures systemic immune states and may not fully represent tissue responses, such as HIV-specific CD8^+^ T cells concentrated in lymph nodes^80^. All participants were assigned male sex at birth, which reflects the burden of HIV among people who use methamphetamine, and all participants with HIV were on ART with suppressed viral load. These features limit confounding by sex and by ongoing viral replication, but they also mean that the findings may not extend to women or to PWH who are not virally suppressed. Nicotine, alcohol and marijuana use were permitted in all groups and are more common among people who use stimulants, so the changes we attribute to methamphetamine use may reflect a broader pattern of polysubstance use.

In summary, methamphetamine use in PWH is associated with persistent CD4^+^ and γδ T cell activation, a lineage-specific shift of monocyte interferon and antigen presentation programs and of CD8^+^ T cell interferon, IL-2 and MYC programs, reduced type 1 effector programming in central memory CD8^+^ T cells, and elevated plasma markers of cytotoxic lymphocyte activation, CXCL13 and neuroaxonal injury. These data identify candidate immune pathways, including TNF, IL-6, IFN-γ and CXCL13 signaling and monocyte antigen presentation to CD8^+^ T cells, that are detectable in blood and that can be tested mechanistically in models of HIV and methamphetamine exposure. Multi-compartment profiling of this kind may help identify biomarkers and therapeutic targets for neuroimmune dysfunction in PWH who use stimulants.

## Data Availability

All data produced in the present study are available upon reasonable request to the authors.

## Acknowledgments

This work was supported by NIH grants awarded by NIDA. R01DA061952 & R01DA052827 to Christina Meade, and R61DA066156 & R01DA063642 to Drew Kiraly. Further support was provided by the Wake Forest Center for Addiction Research.

This study is the first manuscript from the Pathways of Reward and Immune Signaling in Meth-HIV Comorbidity (**PRISM**) team at Wake Forest University School of Medicine, and it reflects the combined efforts of investigators across addiction neuroscience, HIV neurobehavioral research, immunology, precision molecular analysis, molecular imaging, and biostatistics.

The authors would like to thank the study participants who contributed time and samples for this study.

## Supplementary figures

**Supplementary Figure S1.**
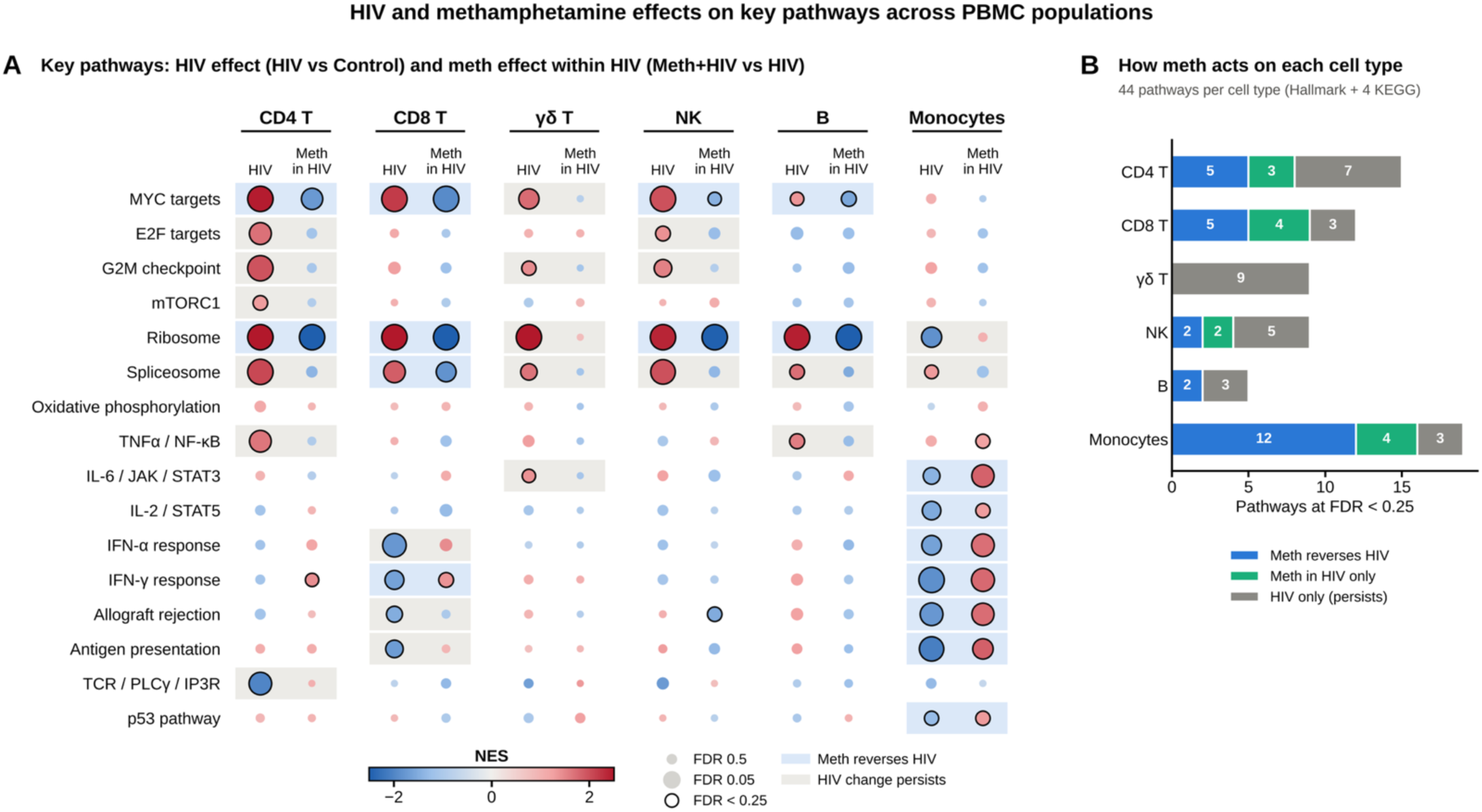
Methamphetamine use opposes HIV-associated pathway changes in monocytes and CD8^+^ T cells but not in CD4^+^ and γδ T cells. Preranked GSEA of pseudobulk expression changes in six PBMC populations (n = 4 per group). Contrasts: HIV, HIV vs Control; Meth in HIV, Meth+HIV vs HIV. **(A)** Normalized enrichment scores (NES) for 16 key Hallmark and KEGG pathways in each cell type. Color, NES (red, higher in the first-named group; blue, lower); size, −log10 FDR; black outline, FDR < 0.25. Blue shading, changed in both contrasts in opposite directions (reversed); gray shading, changed with HIV only (persists). **(B)** Number of the 44 Hallmark and selected KEGG pathways per cell type that were reversed, changed with methamphetamine only or changed with HIV only (FDR < 0.25).

**Supplementary Figure S2.**
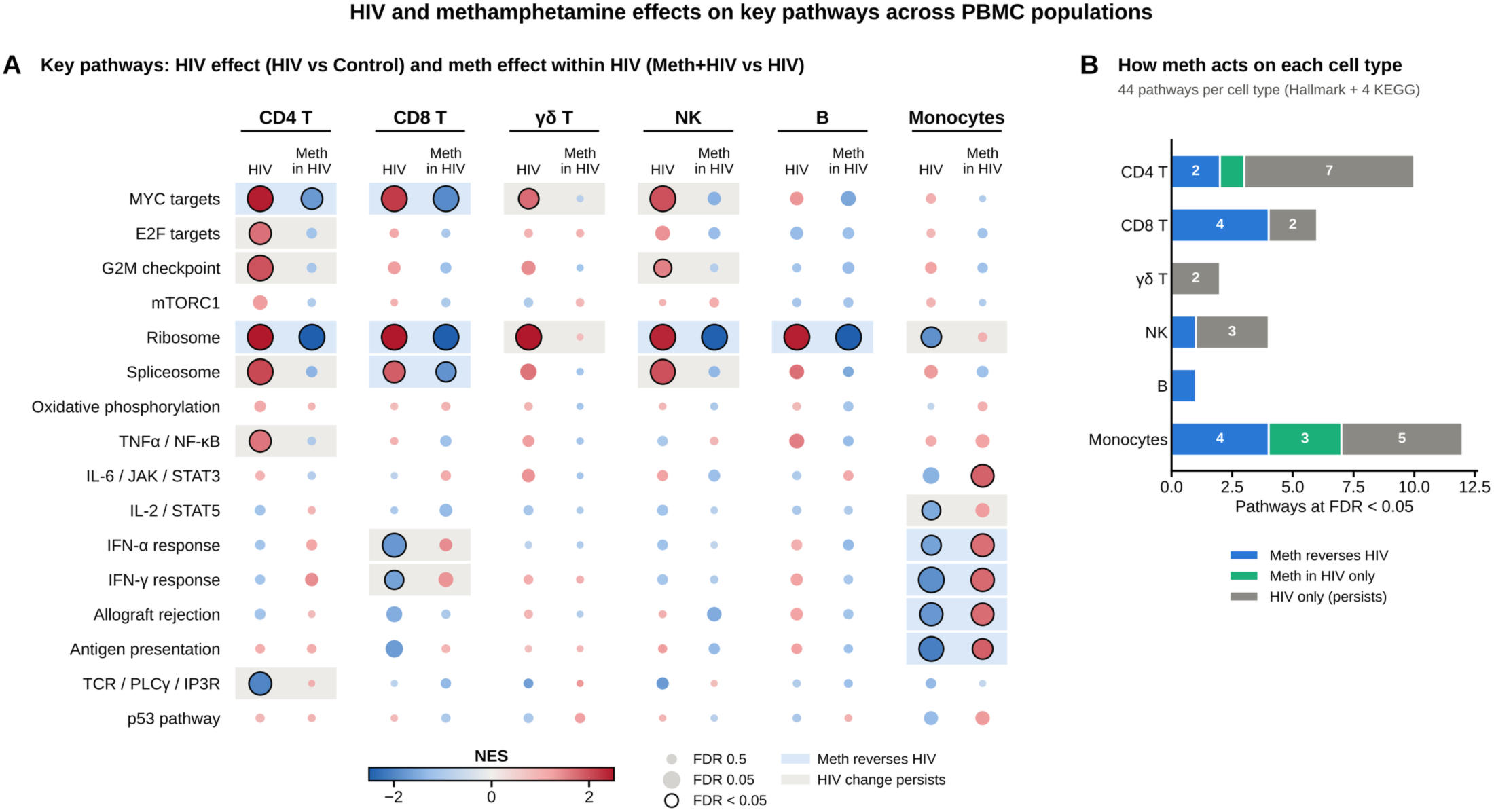
Single-cell pathway classification at FDR < 0.05. Classification of preranked GSEA results at FDR < 0.05. **(A)** NES for 16 representative pathways in each cell type for the HIV (left) and Meth in HIV (right) contrasts. Color, NES (red, higher in the first-named group; blue, lower); size, −log10 FDR; black outline, FDR < 0.05; blue shading, reversed; gray shading, HIV change persists. **(B)** Number of the 44 Hallmark and selected KEGG pathways per cell type that were reversed (blue), changed with methamphetamine only (green) or changed with HIV only (gray) at FDR < 0.05.

**Supplementary Figure S3.**
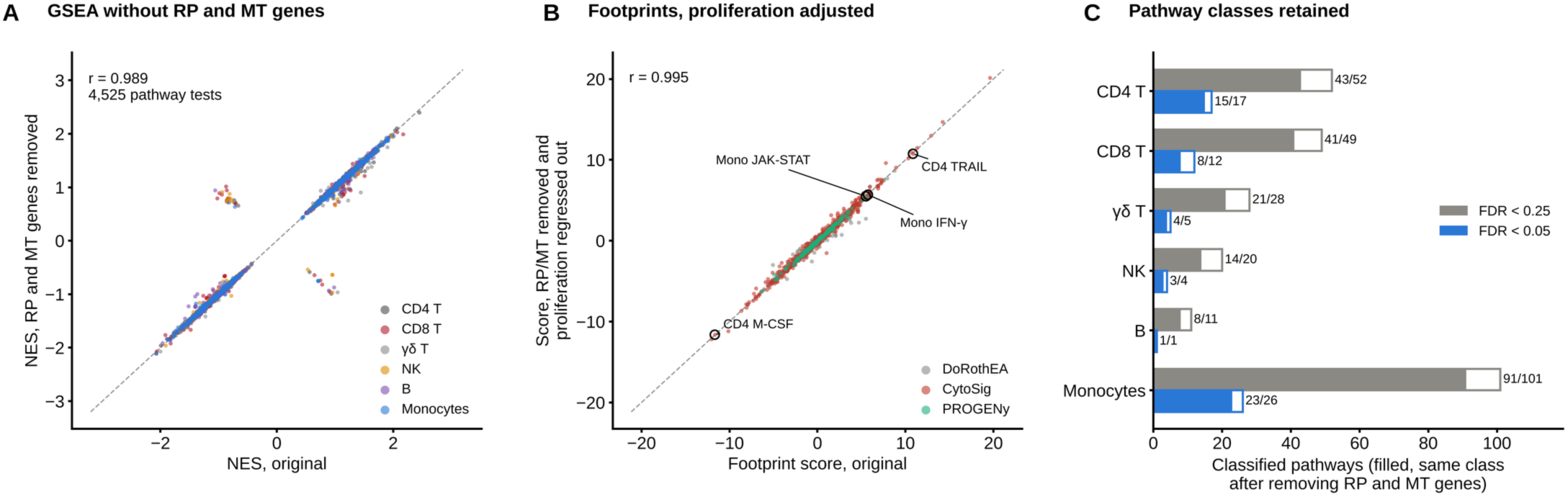
Robustness of single-cell pathway and activity results to ribosomal, mitochondrial and proliferation genes. **(A)** GSEA NES for Hallmark and KEGG gene sets before (x) and after (y) removing ribosomal protein and mitochondrially encoded genes, for HIV vs Control and Meth+HIV vs HIV in each cell type (4,525 tests); dashed line, identity. **(B)** Cytokine response, signaling pathway and transcription factor activity scores before (x) and after (y) removing these genes and adjusting for proliferation genes (G2M checkpoint and E2F targets). Circled points, monocyte IFN-γ response and JAK-STAT activity (Meth+HIV vs HIV) and CD4^+^ T cell TRAIL and M-CSF responses (HIV vs Control). **(C)** Classified pathways per cell type in the original analysis (outline) and those keeping the same class after gene removal (filled), at FDR < 0.25 (gray) and FDR < 0.05 (blue); ribosome and translation sets excluded.

**Supplementary Figure S4.**
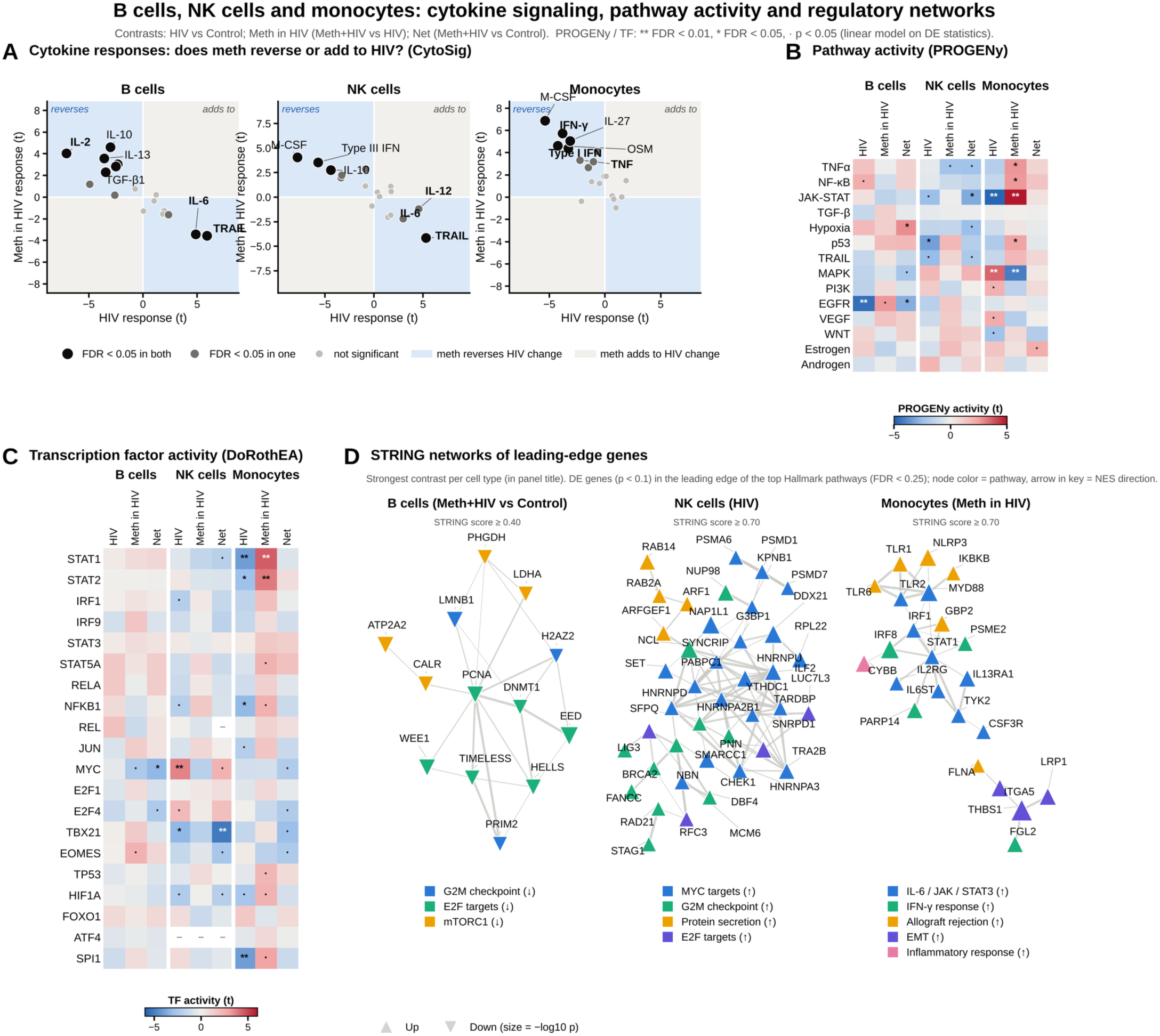
Cytokine response, signaling pathway and transcription factor activity in B cells, NK cells and monocytes. Activities were inferred from the gene-level statistic, sign(log2 fold change) × −log10 p, in B cells, NK cells and monocytes (n = 4 per group). **(A)** CytoSig cytokine response scores (t statistic) in the HIV (x) and Meth in HIV (y) contrasts, for cytokines whose receptor genes were detected (B cells, 17 of 24; NK cells, 17; monocytes, 19). Positive values, stronger response in the first-named group. Blue quadrants, methamphetamine use moves the response opposite to HIV; gray quadrants, same direction. Black, FDR < 0.05 in both contrasts; dark gray, in one; light gray, neither. **(B)** PROGENy activity for 14 signaling pathways in the HIV, Meth in HIV and Net contrasts. **(C)** DoRothEA activity for 20 immune and growth-related transcription factors (regulons A to C); dash, fewer than 10 measured targets. In B and C, red, higher activity in the first-named group; blue, lower; ** FDR < 0.01, * FDR < 0.05, · *p* < 0.05. **(D)** STRING networks built from leading-edge genes of the top Hallmark pathways (FDR < 0.25; gene *p* < 0.1), using the Net contrast for B cells, the HIV contrast for NK cells and the Meth in HIV contrast for monocytes. Node color, pathway; up and down triangles, increased and decreased genes; node size, −log10 p; edge width, STRING score (threshold shown in each panel).

